# CD177, a specific marker of neutrophil activation, is a hallmark of COVID-19 severity and death

**DOI:** 10.1101/2020.12.12.20246934

**Authors:** Yves Lévy, Aurélie Wiedemann, Boris P. Hejblum, Mélany Durand, Cécile Lefebvre, Mathieu Surénaud, Christine Lacabaratz, Matthieu Perreau, Emile Foucat, Marie Déchenaud, Pascaline Tisserand, Fabiola Blengio, Benjamin Hivert, Marine Gautier, Minerva Cervantes-Gonzalez, Delphine Bachelet, Cédric Laouénan, Lila Bouadma, Jean-François Timsit, Yazdan Yazdanpanah, Giuseppe Pantaleo, Hakim Hocini, Rodolphe Thiébaut, the French COVID cohort study group

## Abstract

COVID-19 SARS-CoV-2 infection exhibits wide inter-individual clinical variability, from silent infection to severe disease and death. The identification of high-risk patients is a continuing challenge in routine care. We aimed to identify factors that influence clinical worsening. We analyzed 52 cell populations, 71 analytes, and RNA-seq gene expression in the blood of severe patients from the French COVID cohort upon hospitalization (n = 61). COVID-19 patients showed severe abnormalities of 27 cell populations relative to healthy donors (HDs). Forty-two cytokines, neutrophil chemo-attractants, and inflammatory components were elevated in COVID-19 patients. Supervised gene expression analyses showed differential expression of genes for neutrophil activation, interferon signaling, T- and B-cell receptors, EIF2 signaling, and ICOS-ICOSL pathways in COVID-19 patients. Unsupervised analysis confirmed the prominent role of neutrophil activation, with a high abundance of CD177, a specific neutrophil activation marker. CD177 was the most highly differentially-expressed gene contributing to the clustering of severe patients and its abundance correlated with CD177 protein serum levels. CD177 levels were higher in COVID-19 patients from both the French and “confirmatory” Swiss cohort (n = 203) than in HDs (P< 0.01) and in ICU than non-ICU patients (P< 0.001), correlating with the time to symptoms onset (P = 0.002). Longitudinal measurements showed sustained levels of serum CD177 to discriminate between patients with the worst prognosis, leading to death, and those who recovered (P = 0.01). These results highlight neutrophil activation as a hallmark of severe disease and CD177 assessment as a reliable prognostic marker for routine care.

## Introduction

The Coronavirus Disease 2019 (COVID-19) pandemic is caused by a newly described highly pathogenic beta coronavirus, Severe Acute Respiratory Syndrome coronavirus 2 (SARS-CoV-2) ^1,2^. COVID-19 consists of a spectrum of clinical symptoms that range from mild upper respiratory tract disease in most cases to severe disease that affects approximately 15% of patients requiring hospitalization ^3^, some of whom require intensive care because of severe lower respiratory tract illness, acute respiratory distress syndrome (ARDS), and extra-pulmonary manifestations, leading to multi-organ failure and death. Several recent studies have provided important clues about the physiopathology of COVID-19 ^4-8^. Most compared the immune and inflammatory status of patients at different stages of the disease ^9-11^. Thus, several important biomarkers associated with specific phases of the evolution of COVID-19 have thus far been identified ^12,13^. Inflammation, cytokine storms, and other dysregulated immune responses have been shown to be associated with severe disease pathogenesis ^14,15^. Severe COVID-19 patients are characterized by elevated numbers of monocytes and neutrophils and lymphopenia ^10,16-18^, and a high neutrophil to lymphocyte ratio predicts in-hospital mortality of critically-ill patients ^19^. High levels of pro-inflammatory cytokines, including IL-6,IL-1β, TNF, MCP-1, IP-10, and G-CSF, in the plasma ^5,10,16,18,20^ and a possible defect in type I interferon activity have been reported in patients with severe COVID-19 ^9,15,21^. However, these responses are dynamic, changing rapidly during the clinical course of the disease, which may explain the high variability of the immunological spectrum described ^14,21,22^. This makes it difficult to deduce a unique profile of the pathophysiology of this infection, which is still undetermined. Furthermore, the high amplitude of the signals generated by the inflammation associated with the disease may hide other pathways that are involved.

From a clinical standpoint, clinicians face the daily challenge of predicting worsening patients due to the peculiar clinical course of severe COVID-19, characterized by a sudden deterioration of the clinical condition 7 to 8 days after the onset of symptoms. Determination of the onset of the pathological process once infection has been established in a patient with a severe stage of infection is highly imprecise because of the possible pauci- or asymptomatic phase of the infection, as well as the low specificity of self-limited “flu” illness.

We used a systems immunology approach to identify host factors that were significantly associated with the time to illness onset, severity of the disease (ICU or transfer to ICU), and mortality of COVID-19 patients enrolled in the multicentric French COVID cohort ^23^. In addition to the depletion of T cells and mobilization of B cells, neutrophil activation, and severe inflammation, we show upregulation of CD177 gene expression and protein levels in the blood of COVID-19 patients in both the COVID-19 cohort and a “confirmatory” cohort, i.e., Swiss cohort, relative to healthy subjects. CD177, a neutrophil activation marker, characterized critically ill patients and marked disease progression and death. Our finding highlights the major role of neutrophil activation through CD177 over-expression in the critical clinical transition point in the trajectory of COVID-19 patients.

## Results

### Overview of the phenotype, cytokine, and inflammatory profiles of COVID-19 patients

Patient characteristics from the French COVID cohort enrolled in this analysis are shown in Table 1. All patients from this cohort were stratified as severe according to criteria of the French COVID cohort (clinicaltrials.gov NCT04262921) ^23^, with 53 (87%) hospitalized in an ICU (either initially or after clinical worsening or death) and eight not. The median age was 60 years ([interquartile range (IQR)], [50-69]) and 80% were male. Sampling for immunological analyses was performed within three days of entry and after a median of 11 days [7-14] after symptoms onset. We first assessed leukocyte profiles by flow cytometry using frozen peripheral blood mononuclear cells (PBMCs) from 50 COVID-19 patients (with available PBMC samples) and 18 healthy donors (HDs) (14 or 15 HDs were used as controls per immune-cell subset).

**Table 1.**
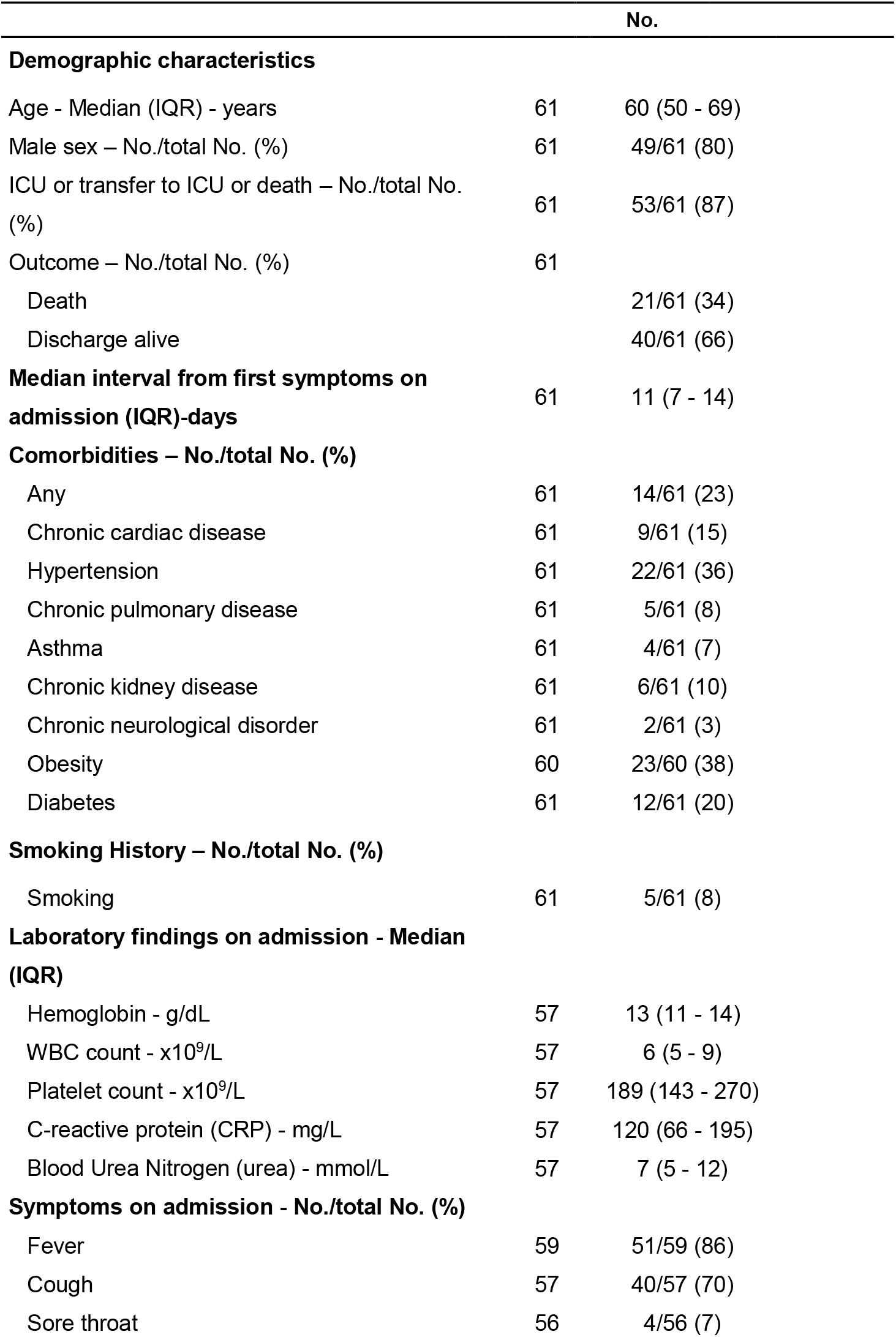
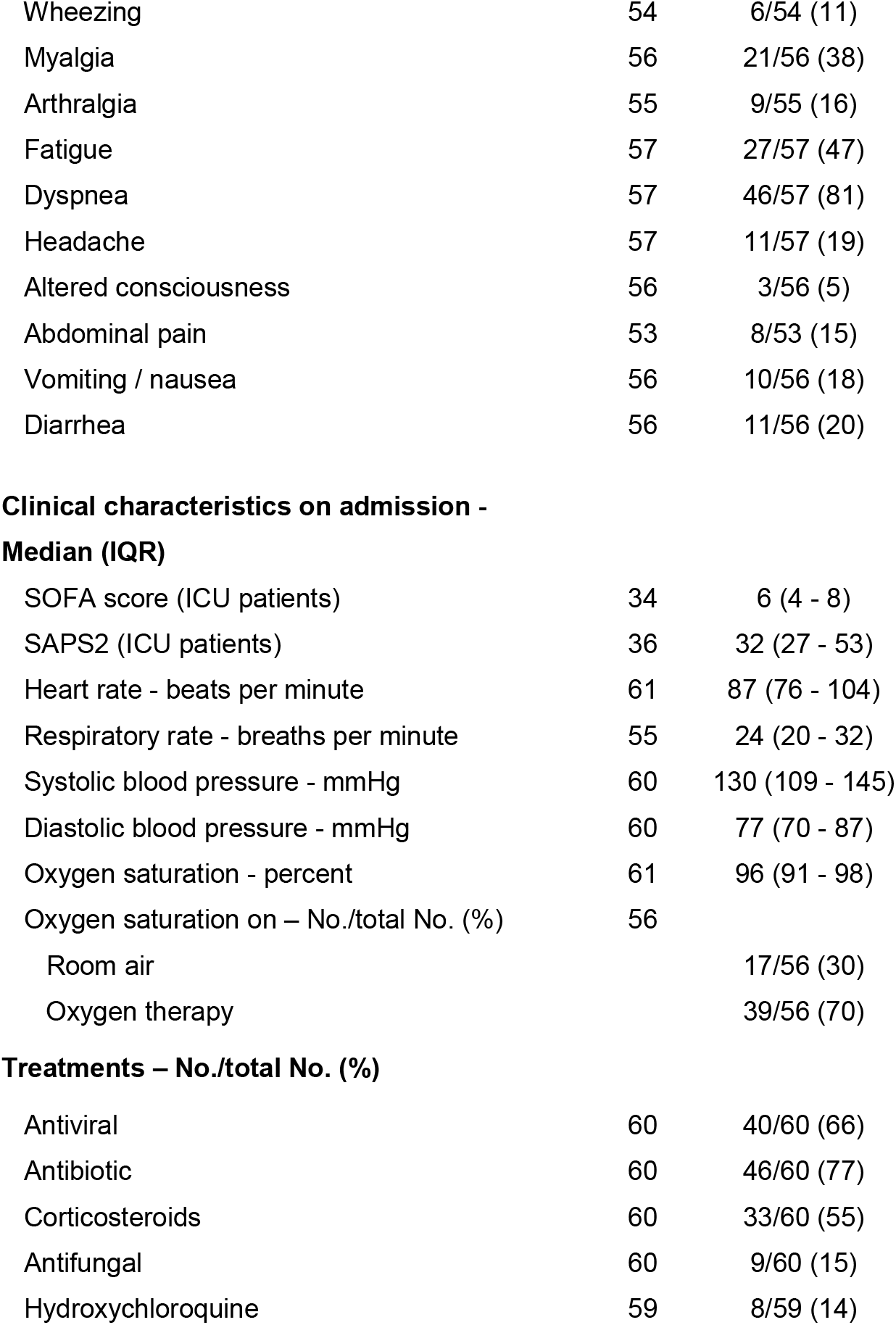
Patient characteristics of the French COVID cohort (n=61)

|  |  | No. |
| --- | --- | --- |
| **Demographic characteristics** |  |  |
| Age - Median (IQR) - years | 61 | 60 (50 - 69) |
| Male sex – No./total No. (%) | 61 | 49/61 (80) |
| ICU or transfer to ICU or death – No./total No. (%) | 61 | 53/61 (87) |
| Outcome – No./total No. (%) | 61 |  |
| Death |  | 21/61 (34) |
| Discharge alive |  | 40/61 (66) |
| **Median interval from first symptoms on admission (IQR)-days** | 61 | 11 (7 - 14) |
| **Comorbidities – No./total No. (%)** |  |  |
| Any | 61 | 14/61 (23) |
| Chronic cardiac disease | 61 | 9/61 (15) |
| Hypertension | 61 | 22/61 (36) |
| Chronic pulmonary disease | 61 | 5/61 (8) |
| Asthma | 61 | 4/61 (7) |
| Chronic kidney disease | 61 | 6/61 (10) |
| Chronic neurological disorder | 61 | 2/61 (3) |
| Obesity | 60 | 23/60 (38) |
| Diabetes | 61 | 12/61 (20) |
| **Smoking History – No./total No. (%)** |  |  |
| Smoking | 61 | 5/61 (8) |
| **Laboratory findings on admission - Median (IQR)** |  |  |
| Hemoglobin - g/dL | 57 | 13 (11 - 14) |
| WBC count - x10<sup>9</sup>/L | 57 | 6 (5 - 9) |
| Platelet count - x10<sup>9</sup>/L | 57 | 189 (143 - 270) |
| C-reactive protein (CRP) - mg/L | 57 | 120 (66 - 195) |
| Blood Urea Nitrogen (urea) - mmol/L | 57 | 7 (5 - 12) |
| **Symptoms on admission - No./total No. (%)** |  |  |
| Fever | 59 | 51/59 (86) |
| Cough | 57 | 40/57 (70) |
| Sore throat | 56 | 4/56 (7) |
| Wheezing | 54 | 6/54 (11) |
| Myalgia | 56 | 21/56 (38) |
| Arthralgia | 55 | 9/55 (16) |
| Fatigue | 57 | 27/57 (47) |
| Dyspnea | 57 | 46/57 (81) |
| Headache | 57 | 11/57 (19) |
| Altered consciousness | 56 | 3/56 (5) |
| Abdominal pain | 53 | 8/53 (15) |
| Vomiting / nausea | 56 | 10/56 (18) |
| Diarrhea | 56 | 11/56 (20) |
| **Median (IQR)** |  |  |
| SOFA score (ICU patients) | 34 | 6 (4 - 8) |
| SAPS2 (ICU patients) | 36 | 32 (27 - 53) |
| Heart rate - beats per minute | 61 | 87 (76 - 104) |
| Respiratory rate - breaths per minute | 55 | 24 (20 - 32) |
| Systolic blood pressure - mmHg | 60 | 130 (109 - 145) |
| Diastolic blood pressure - mmHg | 60 | 77 (70 - 87) |
| Oxygen saturation - percent | 61 | 96 (91 - 98) |
| Oxygen saturation on – No./total No. (%) | 56 |  |
| Room air |  | 17/56 (30) |
| Oxygen therapy |  | 39/56 (70) |
| **Treatments – No./total No. (%)** |  |  |
| Antiviral | 60 | 40/60 (66) |
| Antibiotic | 60 | 46/60 (77) |
| Corticosteroids | 60 | 33/60 (55) |
| Antifungal | 60 | 9/60 (15) |
| Hydroxychloroquine | 59 | 8/59 (14) |

We analyzed 52 immune cell populations, of which 23 showed significant differences (Wilcoxon test adjusted for multiple comparisons) between the COVID-19 patients and HDs. We not only confirmed previously reported abnormalities but also revealed new immunological features of COVID-19 patients (supplementary Figure 1). The COVID-19 patients showed a significant reduction in the frequency of total CD3^+^ T cells and CD8^+^ T cells relative to HDs, as previously reported ^24,25^, that expressed an activated phenotype (CD38^+^HLA-DR^+^) (Figure 1A). The COVID-19 patients also showed lower frequencies of resting memory B cells contrasting with higher frequencies of activated memory B cells and exhausted B cells (Figure 1B). As previously reported ^10,22^, the proportion of plasmablasts was markedly higher in COVID-19 patients (median [Q1-Q3]: 10.85% [3-23]) than HDs (0.76% [0.4-0.8]) (P < 0.001). Total NK-cell frequencies, more precisely those of the CD56^bright^ and CD56^dim^CD57^-^ NK-cell subpopulations, were lower than in HDs (P = 0.017, P < 0.001, and P = 0.004, respectively) (Figure 1C), while a higher proportion of these NK cells, as well as NKT cells, were cycling, expressing Ki67 antigen (CD56^bright^: 22% [13-30], CD56^dim^CD57^-^: 16.8% [11.5-27], and NKT: 10% [5.6-18.2]) (P = 0.003, P = 0.004, and P = 0.001 compared to HD) (Figure 1C). In addition, COVID 19-patients showed significantly smaller classical (CD14^+^CD16^-^), intermediate (CD14^+^CD16^+^), and non-classical (CD14^-^ CD16^+^) monocyte subpopulations than HDs (P = 0.013, P = 0.017, P < 0.001, respectively) (Figure 1D). Interestingly, COVID-19 patients tended to exhibit a higher frequency of γδ T cells than HDs (median 10.4% [7.5-16.1] vs 7.3% [6-10] in HDs; P = 0.068) (Figure 1E), with a significant proportion of γδ T cells showing higher expression of the activation marker CD16 (P = 0.01) and lower expression of the inhibitory receptor NKG2A (P < 0.001) than HDs (Figure 1E). Finally, we observed markedly smaller frequencies of dendritic cells (DCs) for all populations studied (pre-DC, plasmacytoid DC (pDC), and conventional DC (cDC1 and cDC2) in COVID-19 patients than in HDs (P < 0.001, for all comparisons) (Figure 1F).

**Figure 1.**
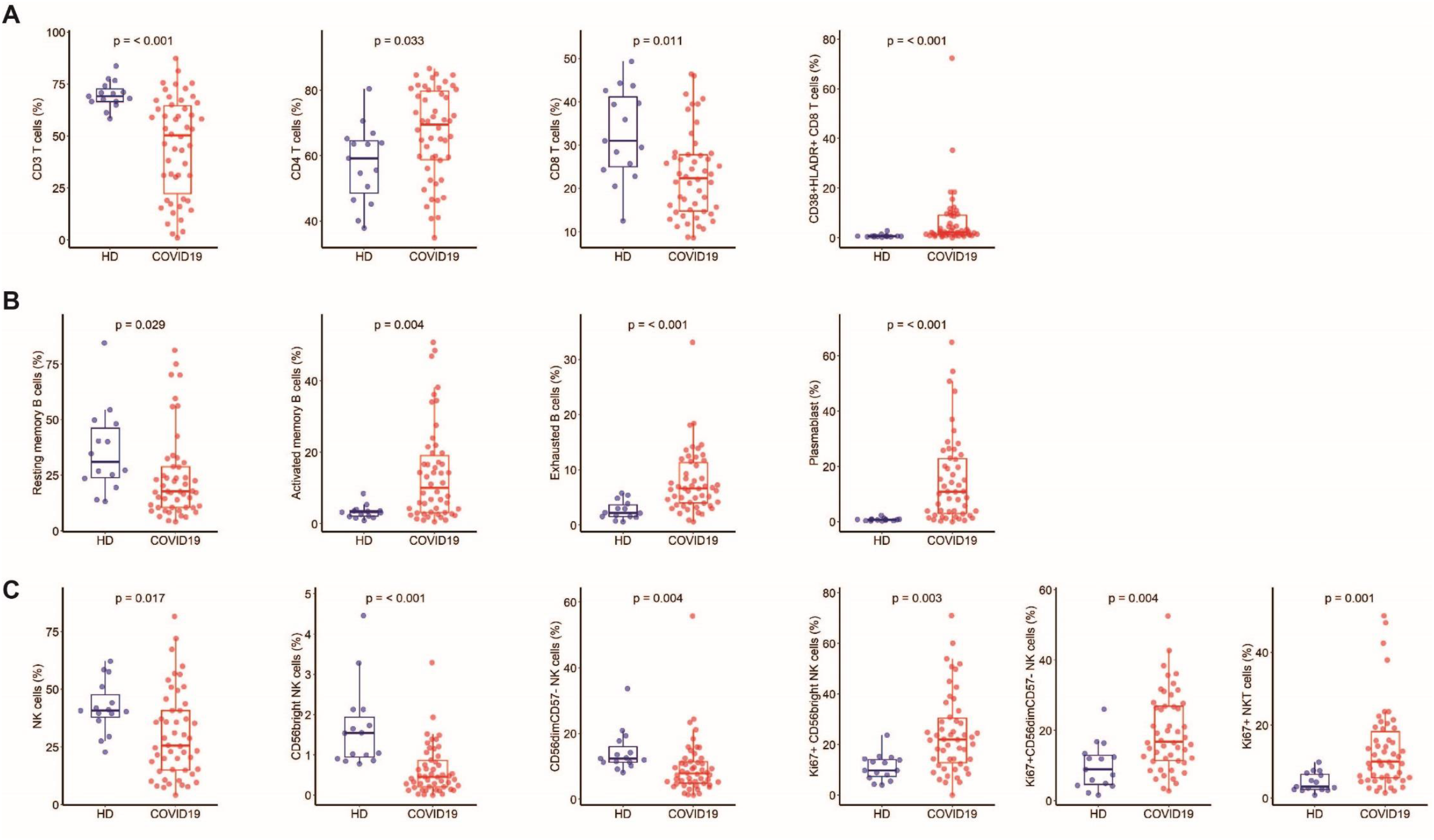
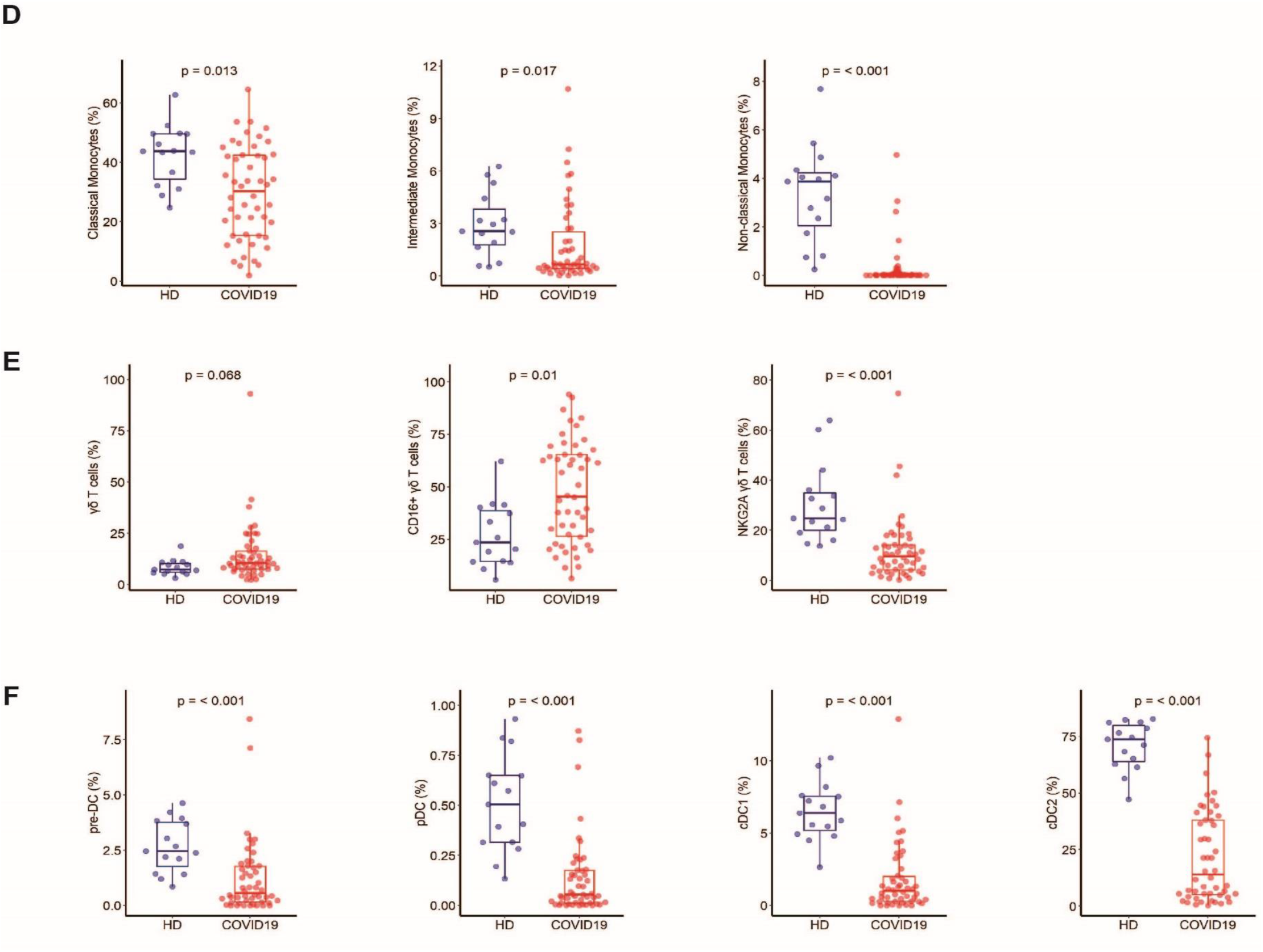
Frequency of immune-cell subsets between HD (n = 18) and COVID-19 patients (n = 50). **A** Frequency of total CD3 T cells, CD4 and CD8 T-cell subsets, and activated CD38^+^HLADR^+^ CD8 T cells. **B** Frequency of B-cell subsets (CD21^+^CD27^+^: resting memory, CD21^-^CD27^+^: activated memory, CD21^-^CD27^-^: exhausted) and plasmablasts (CD38^++^CD27^+^) gated on CD19^+^ B cells. **C** Frequency of NK-cell subsets (gated on CD3^-^ CD14^-^) CD56^Bright^: CD56^++^CD16^+^, CD56^dim^: CD56^+^CD16^++^CD57^+/-^, differentiated Ki67^+^ NK cells (gated on CD56^Bright^ or CD56^dim^CD57^-^ NK cells) and differentiated Ki67^+^ NKT cells (gated on CD3^+^CD56^+^ cells). **D** Monocyte subsets (gated on CD3^-^CD56^-^) (classical monocytes: CD14^+^CD16^-^, intermediate monocytes: CD16^+^CD14^+^, non-classical monocytes: CD14^-^CD16^+^). **E** Frequency of γδ T cells (gated on CD3^+^ T cells) and CD16 and NKG2A expression (gated on γδ CD3 T cells). **F** Frequency of DC subsets (gated on HLADR^+^Lin^-^) (pDC: CD45RA^+^CD33^-^CD123^+^, pre-DC: CD123^+^CD45RA^+^, cDC1: CD33^+^CD123^-^CD141^+^CD1c^low^, cDC2: CD33^+^CD123^-^CD14^+^CD1c^+^) detected by flow cytometry in PBMCs from n = 50 COVID19 patients and n = 18 HDs. The differences between the two groups were evaluated using Wilcoxon rank sum statistical tests. The lower and upper boundaries of the box represent the 25% and 75% percentiles, the whiskers extend to the most extreme data point that is no more than 1.5 times the interquartile range away from the box. Median values (horizontal line in the boxplot) are shown.

We then evaluated the levels of 71 serum cytokines, chemokines, and inflammatory factors in 33 COVID-19 patients and 5 HDs. Forty-four analytes differed significantly (Wilcoxon test adjusted for multiple comparisons) between the COVID-19 patients and HDs (shown in the heatmap in Figure 2 and detailed in Supplementary Figure 2). The levels of 42 factors were higher, among them, pro-inflammatory factors (IL-1a, IL-6, IL-18, tumor necrosis factor-α and β (TNF-α, TNF-β), IL-1ra, ST2/IL-1R4, the acute phase protein lipopolysaccharide binding protein LBP, IFN-a2); Th1 pathway factors (IL-12 (p70), IFN-γ, IP-10, IL-2Ra); Th2/regulatory cytokines (IL-4, IL-10, IL-13); IL-17, which also promotes granulocyte-colony stimulating factor (G-CSF)-mediated granulopoiesis and recruits neutrophils to inflammatory sites; T-cell proliferation and activation factors (IL-7, IL-15); growth factors (SCF, SCGF-b, HGF, b-FGF, b-NGF); and a significant number of cytokines and chemokines involved in macrophage and neutrophil activation and chemotaxis (RANTES (CCL5), MIP-1a and b (CCL3 and CCL4), MCP-1 (CCL2), MCP-3 (CCL7), M-CSF, MIF, Gro-a (CXCL1), monokine inducible by γ interferon MIG/CXCL9, IL-8, IL-9). Interestingly, we found higher levels of midkine, a marker usually not detectable in the serum, which enhances the recruitment and migration of inflammatory cells and contributes to tissue damage ^26^. In parallel, Granzyme B and IL-21 levels were significantly lower in COVID-19 patients than HDs (P = 0.007 and P = 0.004, respectively) (Supplementary Figure 2).

**Figure 2.**
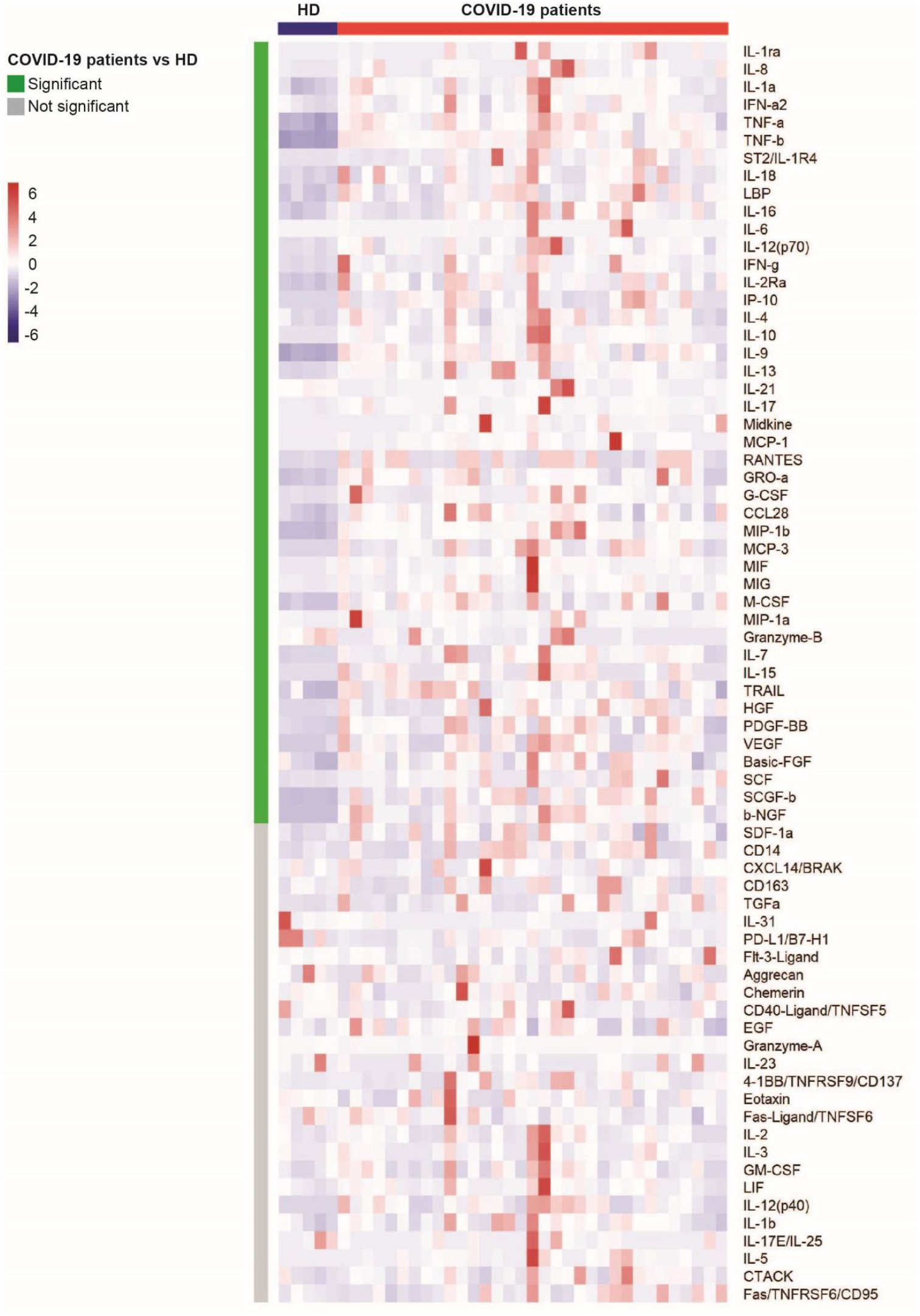
Heatmap of analyte abundance in serum. The colors represent standardized expression values centered around the mean, with variance equal to 1. HD: healthy donors (n = 5), COVID: COVID19 patients (n = 33). Each column represents a subject. Each line represents an analyte.

### Whole blood gene expression profiles show a specific signature for COVID-19 patients

The comparison of gene abundance in whole blood between COVID-19 patients (n = 44) and HDs (n = 10) showed 4,079 differentially expressed genes (DEG) with an absolute fold change ≥ 1.5, including 1,904 that were upregulated and 2,175 that were downregulated (Figure 3A). The main pathways associated with the DEG correspond to the immune response, including neutrophil and interferon signaling, T and B cell receptor responses, metabolism, protein synthesis, and regulators of the EiF2 and mammalian target of rapamycin (mTOR) signaling pathways (Figure 3A). Although several of these pathways involved multiple cell types, analysis of the neutrophil pathway showed higher abundance of genes mainly related to neutrophil activation, their interaction with endothelial cells, and migration (Figure 3B). Among the most highly expressed genes, this signature included CD177, a specific marker of neutrophil adhesion to the endothelium and transmigration ^27^, HP (Haptoglobin), a marker of granulocyte differentiation and released by neutrophils in response to activation ^28^, VNN1 (hematopoietic cell trafficking), GPR84 (neutrophil chemotaxis), MMP9 (neutrophil activation and migration), and S100A8 and S100A12 (neutrophil recruitment, chemotaxis, and migration). The S100A12 protein is produced predominantly by neutrophils and is involved in inflammation and the upregulation of vascular endothelial cell adhesion molecules ^29^ (Figure 3B).

**Figure 3.**
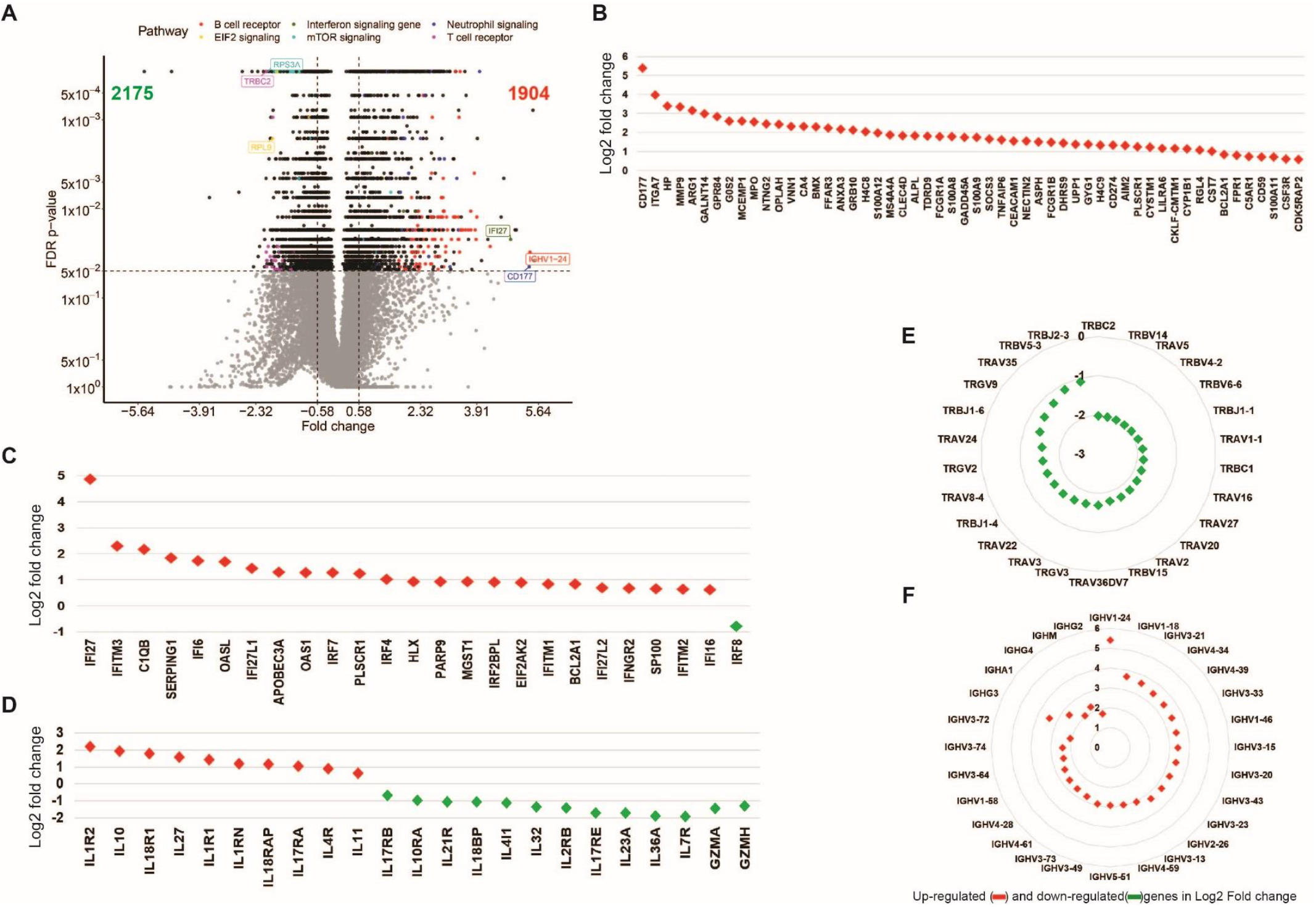
Whole blood gene expression in COVID-19 patients and HDs. **A**. Volcano plot showing differentially expressed genes (DEG) according to the log2 fold change (log2 FC) and Benjamini-Hocberg False Discovery Rate (FDR) with thresholds at absolute log2 FC ≥ log2(1.5) and FDR ≤ 0.05. **B** Main top DEG related to neutrophils. **C** and **D** Main DEG related to IFN and interleukin responses, respectively. **E** Main TCRV T-cell repertoire DEG **F** Main B-cell IGHV repertoire DEG. Red symbols represent overabundant genes in COVID-19 relative to HD, green symbols represent underabundant genes.

In parallel, we observed a higher abundance of several interferon stimulating genes (ISG) (IFI27, IFITM3, IFITM1, IFITM2, IFI6, IRF7, IRF4) (Figure 3C) and cytokines and cytokine receptors (IL-1R, IL-18R1, IL-18RAP, IL-4R, IL-17R, IL-10) (Figure 3D). Consistent with profound T-cell lymphopenia, the expression of several families of T-cell Receptor (TCRA) genes was lower (Figure 3E). We observed severe dysregulation of T-cell function that involved inhibition of serine/threonine kinase PKCθ signaling (z-score = -4.46) (data not shown), as well as the inducible T-cell co-stimulator/ICOSL axis (z-score = -4.5) (Supplementary Figure 3). In contrast to the results for T cells, the peripheral expansion of memory B cells and plasmablasts was associated with broad expansion of the B-Cell Receptor (BCR) (Figure 3F and Supplementary Table 2).

**Table 2.**
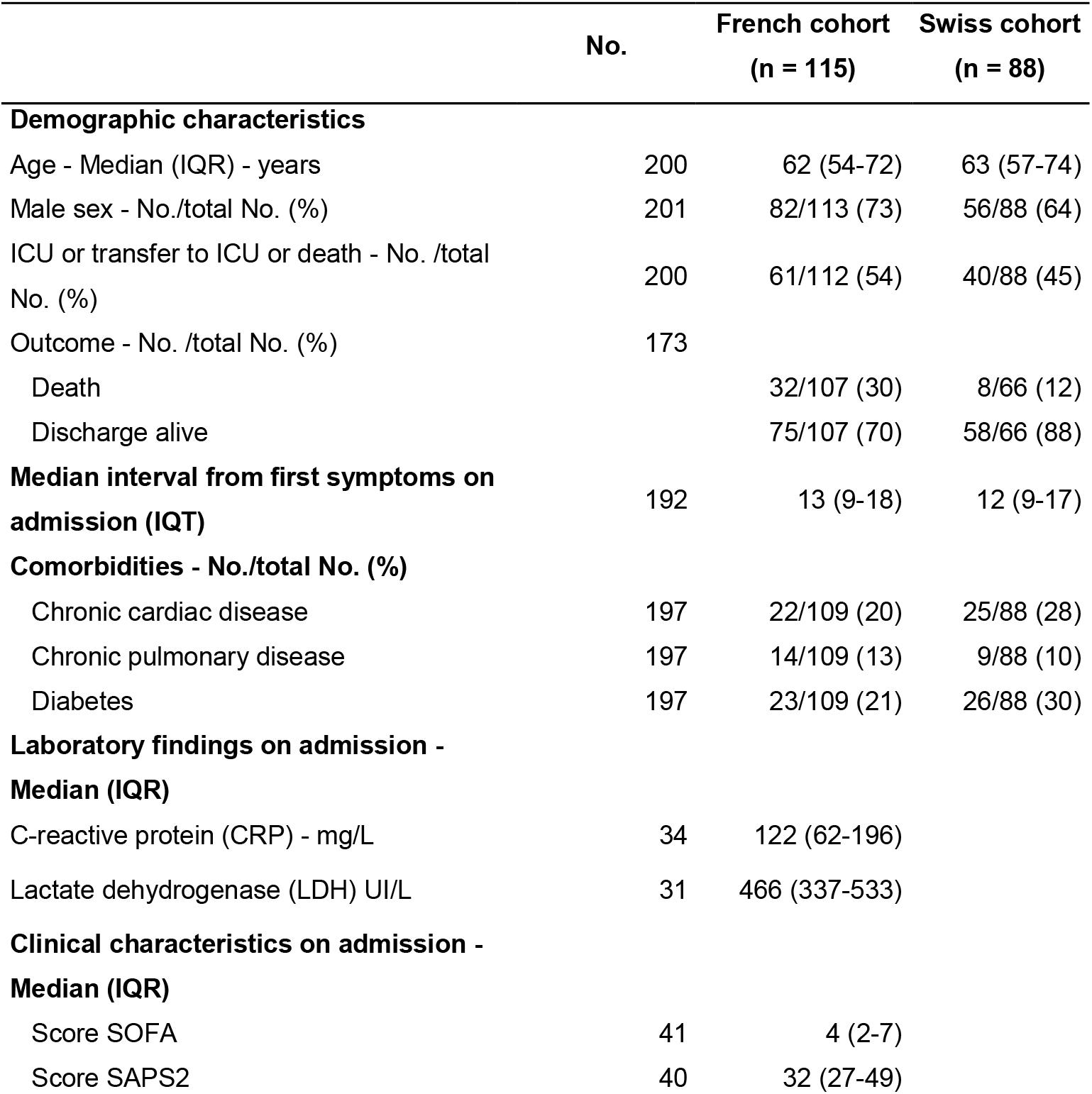
Characteristics of patients involved in the CD177 analysis

|  | No. | French cohort<br>(n = 115) | Swiss cohort<br>(n = 88) |
| --- | --- | --- | --- |
| **Demographic characteristics** |  |  |  |
| Age - Median (IQR) - years | 200 | 62 (54-72) | 63 (57-74) |
| Male sex - No./total No. (%) | 201 | 82/113 (73) | 56/88 (64) |
| ICU or transfer to ICU or death - No. /total No. (%) | 200 | 61/112 (54) | 40/88 (45) |
| Outcome - No. /total No. (%) | 173 |  |  |
| Death |  | 32/107 (30) | 8/66 (12) |
| Discharge alive |  | 75/107 (70) | 58/66 (88) |
| **Median interval from first symptoms on admission (IQT)** | 192 | 13 (9-18) | 12 (9-17) |
| **Comorbidities - No./total No. (%)** |  |  |  |
| Chronic cardiac disease | 197 | 22/109 (20) | 25/88 (28) |
| Chronic pulmonary disease | 197 | 14/109 (13) | 9/88 (10) |
| Diabetes | 197 | 23/109 (21) | 26/88 (30) |
| **Laboratory findings on admission - Median (IQR)** |  |  |  |
| C-reactive protein (CRP) - mg/L | 34 | 122 (62-196) |  |
| Lactate dehydrogenase (LDH) UI/L | 31 | 466 (337-533) |  |
| **Clinical characteristics on admission - Median (IQR)** |  |  |  |
| Score SOFA | 41 | 4 (2-7) |  |
| Score SAPS2 | 40 | 32 (27-49) |  |

We also observed genes belonging to several crucial pathways and biological processes that had not been previously reported to characterize COVID-19 patients to be underrepresented. These included EiF2 signaling, with many downregulated genes, such as ribosomal proteins (RP) and eukaryotic translation initiation factors (EIFs) (Supplementary Figure 3A), common targets of the integrated stress response (ISR), including antiviral defense ^30,31^. In addition, we also found genes involved in signaling through mTOR (supplementary Figure 3B), a member of the phosphatidylinositol 3-kinase-related kinase family of protein kinases. Prediction analysis using Ingenuity pathways showed both lower EIF2 (z-score = - 6.8) and mTOR (z-score = -2.2) signaling in COVID-19 patients than HDs.

### Unsupervised whole blood gene expression profiles reveal distinct features of COVID-19 patients

Unsupervised classification of 44 COVID-19 patients and 10 HDs identified three distinct groups of COVID-19 patients: 10 in group 1, 16 in group 2, and 18 in group 3 (Figure 4). Detailed patient characteristics according to group are presented in supplementary Table 1. Among a large set of clinical and biological characteristics, the analysis showed the differential clustering to not be explained by the severity of the disease. Indeed, the median Sequential Organ Failure Assessment (SOFA) score and Simplified Acute Physiology Score (SAPS2), which include a large number of physiological variables ^32,33^ and evaluate the clinical severity of the disease (a high score is associated with a worse prognosis) of the COVID-19 patients, were 6 [4-7] and 36 [28-53], respectively, with no significant differences between groups. Nevertheless, we observed a significant difference in the median time to symptoms onset at admission, which ranged from 7 [6 –11] days for patients in group 1 to 11 [10-14] and 13 [9-14] days for patients in groups 2 and 3, respectively (P = 0.04, Kruskal-Wallis test). Finally, group 1 which was the closest to HDs in terms of gene profile, consisted of patients in the early days of the disease (Supplementary Table 1).

**Figure 4.**
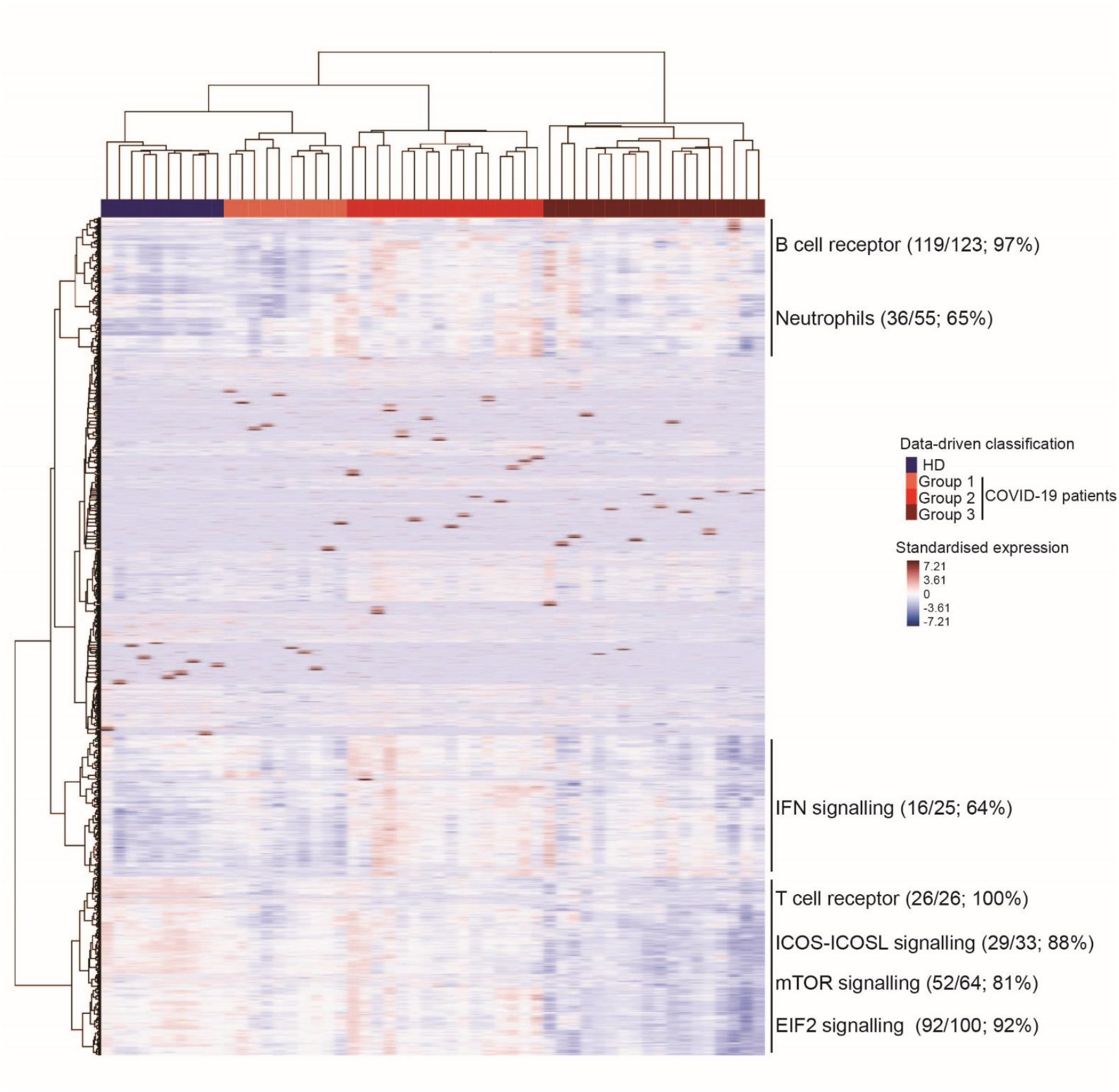
Heatmap of standardized gene expression. The colors represent standardized expression values centered around 0, with variance equal to 1. Each column represents a subject. This heatmap was built by unsupervised hierarchical clustering of log2-counts-per-million RNA-seq transcriptomic data from whole blood (29,302 genes) and subjects (n = 54) using the Euclidean distance and Ward’s method. Seven blocks are highlighted according to the features of gene expression across the groups of individuals. Enrichment (number and % of genes of a given pathway selected in the block) of pathways of interest are shown for each block.

Analysis of the genes contributing to the differences between groups confirmed and extended the findings described above (Figure 3 and Supplementary Figure 3). Several pathways were highly represented in sectors of the heatmap defined according to gene abundance across patient groups. For example, 97% of the genes making up the BCR and 65% of those involved in neutrophil responses were represented among the genes showing a greater abundance in COVID-19 groups 2 and 3 than group 1 and HDs (Figure 4). Other pathways, such as those for interferon (64%), TCR (100%), iCOS-iCOS-L (88%), mTOR (81%), and EiF2 signaling (92%) were also highly represented. The interferon signaling genes, such as IFI44L, IFIT2, and IRF8, a regulator of type I Interferon (α, β), were significantly more abundant at earlier stages (in patients from group 2) and tended to be less abundant in group 3, at more advanced stages of the disease. Finally, the abundance of genes belonging to T-cell pathways (TCR, iCOS-iCOSL signaling) or mTOR and EIF2 signaling was lower in group 3, that is to say, those who were analyzed after a longer time to symptoms onset. The findings described above highlight the heterogeneity of COVID-19 patients.

### Integrative analysis of all biomarkers reveals the major contribution of CD177 in the clinical outcome of COVID-19 patients

We performed an integrative analysis using all available data to disentangle the relative contribution of the various markers at the scale of every patient. We thus pooled the data for 29,302 genes from whole blood RNA-seq, cell phenotypes (52 types), and cytokines (71 analytes) using the recently described MOFA approach ^34^, which is a statistical framework for dimension reduction adapted to the multi-omics context. The data are reduced to components that are linear combinations of variables explaining inter-patient variability across the three biological measurement modalities. The first component, that we called our integrative score, discriminated between the three groups of COVID-19 patients and HDs (Figure 5A), although it only explained a portion of the variability within each of the three types of markers (14% of gene expression, 14% of cell phenotypes, and 5% of cytokines). The main contributors for the cell phenotype were the significantly lower frequency of cDC2 and T cells and, marginally, the higher number of plasmablasts and CD16^+^ γδ T cells in COVID-19 patients (Figure 5B). The contribution of soluble factors was marked by higher levels of soluble CD163 (sCD163), a marker of polarized M2 macrophages involved in tissue repair ^35^, in more advanced COVID-19 groups (Figure 5C). Indeed, CD163 gene expression was also significantly higher in COVID-19 patients than HDs (log2 fold change = +1.55; FDR = 4.79 10^−2^). sCD163 has also been reported to be a marker of disease severity in critically ill patients with various inflammatory or infectious conditions ^36^. Interestingly, the genes that contribute the most to the synthesis of this factor were part of the neutrophil module (CD177, ARG1, MMP9) (Figure 5D). The increasing abundance of CD177 gene expression according to group was again clearly apparent (Supplementary Figure 4). Integrated analysis also revealed higher expression of proprotein convertase subtilisin/kexin type 9 (PCSK9). High plasma PCSK9 levels highly correlate with the development and aggravation of subsequent multiple organ failure during sepsis ^37,38^. Of note, high PCSK9 levels have been recently associated with severe Dengue infection ^39^.

**Figure 5.**
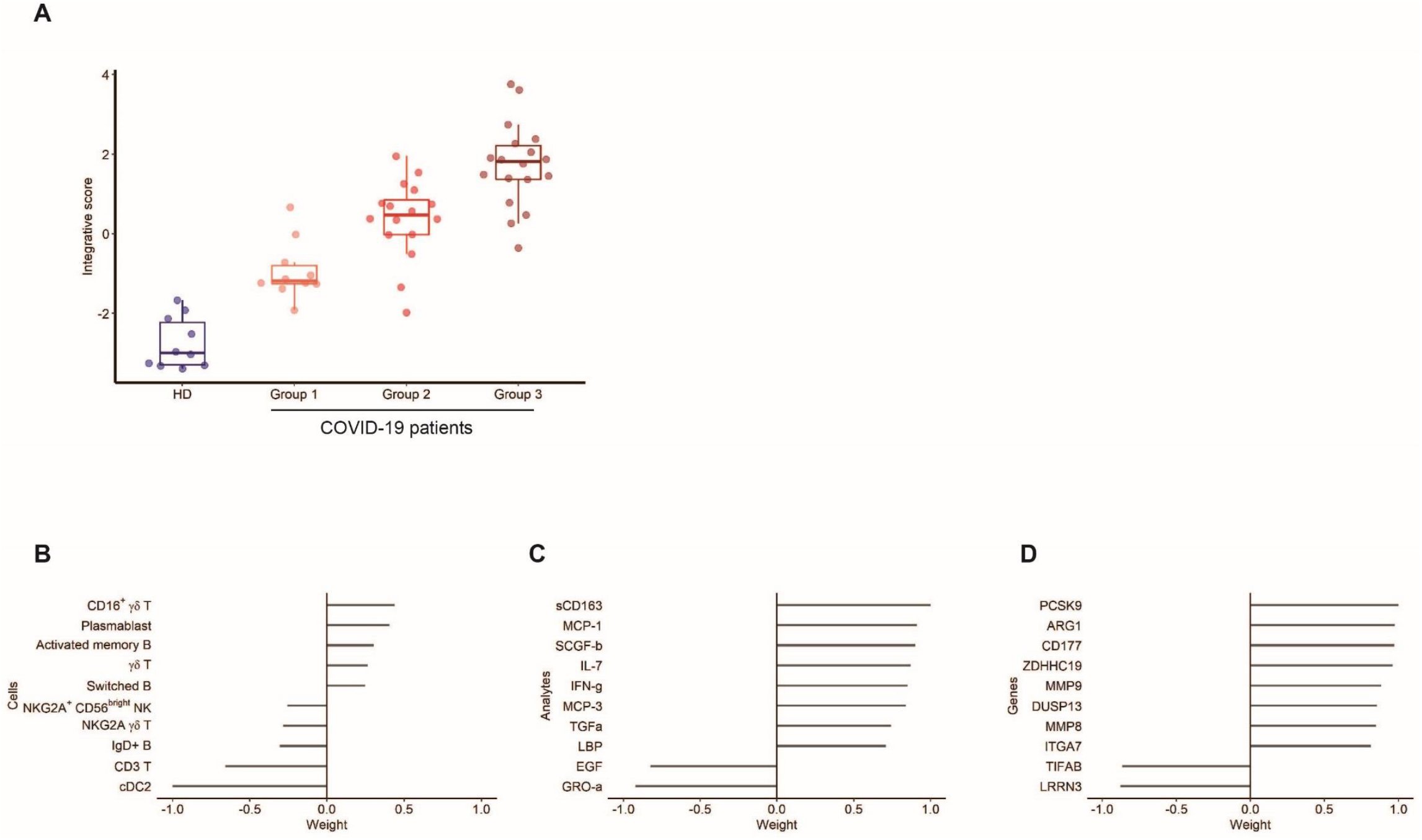
Integrative analysis. Integrative analysis of the data of RNA-seq (29,302 genes) from 44 COVID-19 patients, cell phenotype (52 types) from 45 COVID-19 patients, and serum analytes (71 analytes) from 33 COVID-19 patients using a sparse principal component analysis approach, MOFA v2. **A** Integrative score according to the patient groups defined by the hierarchical clustering of the RNA-seq data. Top 10 marker contributions (according to the weight from -1 to 1) of the cell phenotypes (**B**), serum analytes (**C**), and RNA-seq (**D**). The integrative score corresponds to the first factor of the analysis and allows the ordering of individuals along an axis centered at 0. Individuals with an opposite sign for the factor therefore have opposite characteristics.

### Serum CD177 protein levels are associated with the clinical outcome of COVID-19 patients

Given the contribution of the neutrophil activation pathway in the clustering of COVID-19 patients, we sought neutrophil-activation features that could act as possible reliable markers of disease evolution. We focused on CD177 because: i) it is a neutrophil-specific marker representative of neutrophil activation, ii) it was the most highly differentially expressed gene in patients, and iii) the protein can be measured in the serum, making its use as a marker clinically applicable. Thus, we used an ELISA to quantify CD177 in the serum of 203 COVID-19 patients (115 patients from the French cohort and 88 patients from the Swiss COVID-19 cohort that we used as “a confirmatory” cohort, patient characteristics are described in Table 2), 21% of whom the measurements were repeated (from 2 to 10 measurements per individual). First, we confirmed the significantly higher median serum protein level in the global cohort of COVID-19 patients (4.5; [2.2-7.4]) relative to that of 16 HDs (2.2 [0.9-4.2]) (P = 0.015, Wilcoxon test) (Figure 6A). Second, we found a robust agreement between CD177 gene expression measured by RNA-seq and CD177 protein levels measured by ELISA (intraclass correlation coefficient 0.88), (Figure 6B).

**Figure 6:**
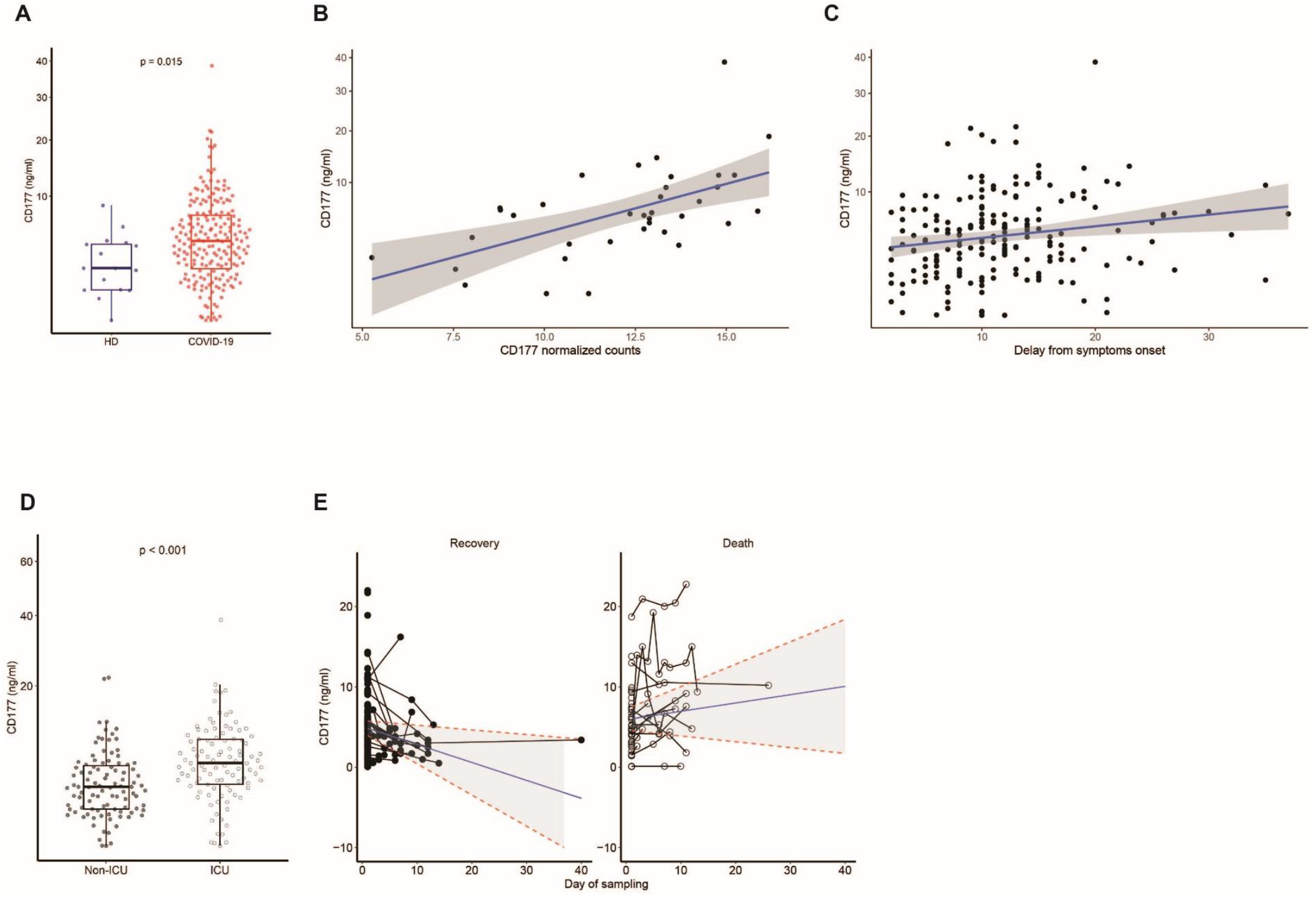
Distribution of the CD177 marker and association with clinical outcomes of COVID-19 patients. **A** Measurement of CD177 (ng/ml). HD: Healthy donors (n = 16), COVID-19 patients (n = 203). The difference between the two groups was evaluated using Wilcoxon rank sum statistical tests. The median values (horizontal line in the boxplot) are shown. The lower and upper boundaries of the box represent the 25% and 75% percentiles. **B** Correlation between normalized CD177 values of gene expression measured by RNA-seq and CD177 protein by ELISA (ng/ml) from 36 COVID-19 patients. The blue line represents the linear regression line and the grey area the 95% prediction confidence interval. **C** Association between CD177 serum concentration and time from symptoms onset (n = 192). This association was tested using Spearman correlation tests. The blue line represents the linear regression line and the grey area the 95% confidence interval. **D** Measurement of CD177 serum concentration in patients hospitalized in an intensive care unit (ICU) or not (n = 196). Wilcoxon rank tests were used. The median values (horizontal line in the boxplot) are shown. The lower and upper boundaries of the box represent the 25% and 75% percentiles. **E**. Change of CD177 concentration over time according to the occurrence of death for 172 COVID-19 patients and a total of 248 measurements. Predictions were calculated using a mixed effect models for longitudinal data.

Then, we examined the association of clinical characteristics and outcomes with serum CD177 concentration at the time of admission. The serum CD177 concentration was positively associated with the time to symptoms onset (P = 0.0026) (Figure 6C) and was higher for patients hospitalized in an ICU (6.0 ng/ml [3.5-9.4] vs 3.3 ng/ml [1.5-5.6], P < 0.001) (Figure 6D). The association between serum CD177 levels and hospitalization in an ICU was independent of the usual risk factors, such as age, sex, chronic cardiac or pulmonary diseases, or diabetes (multivariable logistic regression, adjusted odds ratio 1.14 per unit increase, P < 0.001). We observed a trend towards a positive association with the SOFA and SAPS2 risk scores that was not statistically significant (P = 0.17 and P = 0.074, respectively) (supplementary Figure 5A and B). CD177 levels were not associated with other conditions that contribute to a high risk of severe disease, such as diabetes (P = 0.632), chronic cardiac disease (P = 0.833), chronic pulmonary disease (P = 0.478), or age of the patient (P = 0.83) (data not shown).

We then examined the dynamics of the CD177 concentration in 172 COVID-19 patients, with longitudinal serum samples, using all available measurements (Figure 6E). At the first measurement, the average concentration of CD177 was not significantly different between the patients who died and those who recovered (5.93 vs 5.06, P= 0.26, Wald test). However, CD177 levels decreased significantly in those who recovered (−0.22 ng/mL/day, 95% CI - 0.307; -0.139), whereas it was stable in those who died (+0.10 ng/mL/day, 95% CI +0.014; +0.192) and the difference between the two groups was statistically significant (Wald test for interaction, P = 0.010). These results show that the stability of CD177 protein levels in severe COVID-19 patients during the course of the disease is a hallmark of a worse prognosis, leading to death.

## Discussion

Here, we investigated factors that influence the clinical outcomes of severe COVID-19 patients involved in a multicentric French cohort combining standardized whole-blood RNA-Seq analyses, in-depth phenotypic analysis of immune cells, and measurements of a large panel of serum analytes. An integrated and global overview of host markers revealed several pathways associated with the course of COVID-19 disease, with a prominent role for neutrophil activation. This signature included CD177, a specific neutrophil marker of activation, adhesion to the endothelium, and transmigration. The correlation between CD177 gene abundance and serum protein levels in the blood of COVID-19 patients underscores the importance of this marker, making the measurement of CD177 levels a reliable approach that is largely accessible in routine care. We also demonstrated that the dynamics of serum CD177 levels is strongly associated with the severity of COVID-19 disease, ICU hospitalization, and survival in an additional cohort of patients.

CD177 is a glycosylphosphatidylinositol (GPI)-anchored protein expressed by a variable proportion of human neutrophils. It plays a key role in the regulation of neutrophils by modulating their migration and activation. For example, the CD177 molecule has been identified as the most dysregulated parameter in purified neutrophils from septic shock patients ^40^ and in severe influenza ^41^. Clinically, neutrophil chemotaxis, infiltration of endothelial cells, and extravasation into alveolar spaces have been described in lung autopsies from deceased COVID-19 patients ^19^. Elevated CD177 mRNA expression has also been described for patients with acute Kawasaki Disease (KD) ^42^ and resistant to IV Ig therapy ^43,44^, a syndrome that has been described as a possible complication of SARS-CoV-2 infection in children ^45,46^. Our results are also consistent with those obtained using animal models, suggesting an important role for neutrophil activation in the severity of infection with respiratory viruses through their migration towards infected lungs, and in humans infected with influenza ^47-49^.

The neutrophil activation signature is a specific feature of the homing of activated neutrophils toward infected lung tissue in acute lung injury ^50^, followed by the initiation of aggressive responses and the release of neutrophil extracellular traps (NETs), leading to an oxidative burst and the initiation of thrombus formation ^51^. Previous studies have reported elevated levels of circulating NETs in COVID-19 disease ^52^. Consistent with this finding and extending these data, we showed the differential expression of NET-related genes ^41,47,48^ (S100A8, S100A9, and S100A12), confirming the recently described elevated expression of calprotectin (heterodimer of S100A8 and A9) in severe COVID-19 patients ^13^. The association of neutrophil activation signature with COVID-19 severity has also been described recently with CD177 gene being one of the most differentially expressed gene in advanced disease ^53^. Although, it is difficult to formally conclude whether CD177 is a causal factor of disease progression or a consequence of the severity of the disease, our data strongly show that CD177 is a valid hallmark of the physiopathology of COVID-19. This observation suggests that the activation of neutrophils, triggered by the infection, is a fundamental element of the innate response. However, persistent activation of this pathway may constitute, along with other factors (e.g., “cytokine storm”), fatal harm possibly associated with the critical turning point in the clinical trajectory of patients during the second week of the infection.

Neutrophil activation was associated with significant changes in the level of gene expression of several pathways, some concordantly associated with disturbances in immune-cell populations and cytokine and inflammatory profiles. We reveal a complex picture of the activation of innate immunity, assessed by significant changes in the expression of several genes involved in interferon signaling and the response to stress and the production of inflammatory/activation markers, with a balance between pro-inflammatory signals (increased expression of IL-1R1, IL-18R1 and its accessory chain IL-18 RAP) and anti-inflammatory cytokines or regulators (increased expression of IL-10, IL-4R, IL-27, IL-1RN) ^54,55^. The frequency of γδ T cells, a subpopulation of CD3^+^ T cells that were first described in the lung ^56^ and that play critical roles in anti-viral immune responses, tissue healing, and epithelial cell maintenance ^57^, was elevated and they expressed an activation marker (CD16) and low levels of the inhibitory receptor NKG2A, suggesting possible killing capacity. In accordance with our observation that EiF2 signaling is significantly inhibited in COVID-19 patients, recent studies have shown that coronaviruses encode ISR antagonists, which act as competitive inhibitors of EiF2 signaling ^58,59^. Similarly, the inhibition of mTOR signaling that we found in COVID-19 patients is consistent with the impaired mTOR signaling reported in blood myeloid dendritic cells of COVID-19 patients ^21^. Interestingly, we observed a markedly smaller proportion of all DC (pre-DC, pDC, cDC1 and cDC2) and monocyte cell populations, including classical, intermediate, and non-classical subpopulations, in COVID-19 patients than in HDs. Based on these observations, it can be hypothesized that the impairment in IFN-α production observed in severe COVID-19 patients may be the result of both a decrease in the number of pDCs, which are natural IFN-producing cells ^60^, and inhibition of mTOR signaling, a regulator of IFN-α production, in these cells ^61^.

We confirmed the previously reported expansion of B-cell populations ^21,22^ in COVID-19 patients and our results showed that the anti-SARS-CoV-2 B cell repertoire is commonly mobilized. The marked upregulation of IgV gene families included the IgHV1-24 family, described to be specific for COVID-19 ^62^. The expanded VH4-39 family was also recently reported to be the most highly represented in S-specific SARS-CoV-2 sequences ^62^. We also found enrichment of VH3-33, previously described in a set of clonally related anti-SARS-CoV-2 receptor-binding domain antibodies^63^.

Globally, these results show that the defense against SARS-CoV-2 following pathogen recognition triggers a fine-tuned program that not only includes the production of antiviral (Interferon signaling) and pro-inflammatory cytokines but also signals the cessation of the response and a strong disturbance of adaptive immunity.

The same pathways (immune and stress responses through EiF2 signaling, neutrophil and Interferon signaling, T- and B-cell receptor responses, and mTOR pathways) contributed to the ability to discriminate between three groups of severe COVID-19 patients in an unsupervised analysis. One limitation of our study was that we did not repeat the RNA-seq analyses in these specific groups of patients. Nonetheless, it is noteworthy that these groups differed significantly by the time from disease onset. These findings provide clues in our understanding of the wide range of profiles previously described for COVID-19 by showing that these patterns may be mainly related to time-dependent changes in the blood during the course of the infection ^15,64^. For example, the observed lower abundance of IFN signaling genes in the last group of COVID-19 patients may be due to decreased abundance in more advanced disease and/or patients who constitutively present a lower abundance of ISG, as described in previous studies ^65^.

Several months after the emergence of this new disease, treatment options for patients with severe disease requiring hospitalization are still limited to corticosteroids, which has emerged as the treatment of choice for critically ill patients ^66,67^. However, interventions that can be administered early during the course of infection to prevent disease progression and longer-term complications are urgently needed. A major obstacle for the design of “adapted” therapies to the various stages of disease evolution is a lack of markers associated with sudden worsening of the disease of patients with moderate to severe disease and markers to predict improvement. Our results show that the measurement of CD177 during the course of the disease may be helpful in following the response to treatment and revision of the prognosis. In addition, they suggest that therapies aiming to control neutrophil activation and chemotaxis should be considered for the treatment of hospitalized patients.

## Materials and Methods

### Subjects

We enrolled a subgroup of COVID19 patients of the prospective French COVID cohort (registered at clinicaltrials.gov NCT04262921) in this immunological study which is part of the cohort main objectives. Ethics approval was given on February 5th by the French Ethics Committee CPP-Ile-de-France VI (ID RCB: 2020-A00256-33). Eligible patients were those who were hospitalized with virologically confirmed COVID-19. Briefly, nasopharyngeal swabs were performed on the day of inclusion for SARS-CoV-2 testing according to WHO or French National Health Agency guidelines. Viral loads were quantified by real-time semi-quantitative reverse transcriptase polymerase chain reactions (RT-PCR) using either the Charité WHO protocol (testing the E gene and RdRp) or the Pasteur institute assay (testing the E gene and two other RdRp targets, IP2 and IP4). The study was conducted with the understanding and the consent of each participant or its surrogate covering the sampling, storage, and use of biological samples. The Swiss cohort was approved by the ethical commission (CER-VD; Swiss ethics protocol ID: 2020-00620) and all subjects provided written informed consent. Blood from healthy donors was collected from the French Blood Donors Organization (Etablissement Français du sang (EFS)) before the COVID-19 outbreak. HD characteristics are shown in Supplementary Table 3.

### Quantification of serum analytes

In total, 71 analytes were quantified in heat-inactivated serum samples by multiplex magnetic bead assays or ELISA. Serum samples from five healthy donors were also assayed as controls. The following kits were used according to the manufacturers’ recommendations: LXSAHM-2 kits for CD163,ST2,CD14 and LBP (R&D Systems); the LXSAHM-19 kit for IL-21, IL-23, IL-31, EGF, Flt-3 Ligand, Granzyme B, Granzyme A, IL-25, PD-L1/B7-H1, TGF-α, Aggrecan, 4-1BB/CD137, Fas, FasL,CCL-28, Chemerin, sCD40L, CXCL14, and Midkine (R&D Systems); and the 48-Plex Bio-Plex Pro Human Cytokine screening kit for IL-1β, IL-1rα, IL-2, IL-4, IL-5, IL-6, IL-7, IL-8 / CXCL8, IL-9, IL-10, IL-12 (p70), IL-13, IL-15, IL-17A / CTLA8, Basic FGF (FGF-2), Eotaxin / CCL11, G-CSF, GM-CSF, IFN-γ, IP-10/CXCL10, MCP-1 /CCL2, MIP-1α / CCL3, MIP-1β / CCL4, PDGF-BB (PDGF-AB/BB), RANTES/CCL5, TNF-α, VEFG (VEGF-A), IL-1a, IL-2Ra (IL-2R), IL-3, IL-12 (p40), IL-16, IL-18, CTACK / CCL27, GRO-a /CXCL1 (GRO), HGF, IFN-α2, LIF, MCP-3 / CCL7, M-CSF, MIF, MIG/CXCL9, b-NGF, SCF, SCGF-b, SDF-1α, TNF-b/LTA, and TRAIL (Bio-Rad). The data were acquired using a Bio-Plex 200 system. Extrapolated concentrations were used and the out-of-range values were entered at the highest or lowest extrapolated concentration. Values were standardized for each cytokine across all displayed samples (centered around the observed mean, with variance equal to 1). CD177 quantification was performed on non-inactivated serum samples (diluted 1:2 or 1:10) using a Human CD177 ELISA Kit (ThermoFisher Scientific), according to the manufacturer’s instructions.

### Cell phenotyping

Immune-cell phenotyping was performed using an LSR Fortessa 4-laser (488, 640, 561, and 405 nm) flow cytometer (BD Biosciences) and Diva software version 6.2. FlowJo software version 9.9.6 (Tree Star Inc.) was used for data analysis. CD4^+^ and CD8^+^ T cells were analyzed for CD45RA and CCR7 expression to identify the naive, memory, and effector cell subsets for co-expression of activation (HLA-DR and CD38) and exhaustion/senescence (CD57and PD1) markers. CD19^+^ B-cell subsets were analyzed for the markers CD21 and CD27. ASC (plasmablasts) were identified as CD19^+^ cells expressing CD38 and CD27. We used CD16, CD56, and Ki57 to identify NK-cell subsets. γδ T cells were identified using an anti-TCR γδ antibody. HLA-DR, CD33, CD45RA, CD123, CD141, and CD1c were used to identify dendritic cell (DC) subsets, as previously described ^68^

### RNA sequencing

Total RNA was purified from whole blood using the Tempus™ Spin RNA Isolation Kit (ThermoFisher Scientific). RNA was quantified using the Quant-iT RiboGreen RNA Assay Kit (Thermo Fisher Scientific) and quality control performed on a Bioanalyzer (Agilent). Globin mRNA was depleted using Globinclear Kit (Invitrogen) prior to mRNA library preparation with the TruSeq® Stranded mRNA Kit, according to the Illumina protocol. Libraries were sequenced on an Illumina HiSeq 2500 V4 system. Sequencing quality control was performed using Sequence Analysis Viewer (SAV). FastQ files were generated on the Illumina BaseSpace Sequence Hub. Transcript reads were aligned to the hg18 human reference genome using Salmon v0.8.2 ^69^ and quantified relative to annotation model “hsapiens_gene_ensembl” recovered from the R package biomaRt v2.42.1 ^70^. Quality control of the alignment was performed via MultiQC v1.4 ^69^. Finally, counts were normalized as counts per million.

### Statistical analysis

Subgroups of COVID-19 patients were identified from unsupervised hierarchical clustering of log2-counts-per-million RNA-seq transcriptomics from whole-blood using the Euclidean distance and Ward’s method. Differential expression analysis was carried out using dearseq ^71^ to contribute to the analysis of genes of which the abundance differed across the three COVID-19 patient subgroups and healthy subjects. Once the groups were defined by hierarchical clustering, the analysis of the genes contributing to each group was performed by selecting genes with an absolute fold-change ≥ 1.5 in the comparison of interest for which the difference in expression between HDs and COVID-19 patients was significant (P ≤ 0.05) (to avoid so called “double dipping” [https://arxiv.org/abs/2012.02936]). Pathway analyses of the genes involved in each comparison was performed using Ingenuity Pathway Analysis (IPA ®, Qiagen, Redwood City, California, Version 57662101, 2020). For canonical pathway analysis, a Z-score ≥ 2 was defined as the threshold for significant activation, whereas a Z-score ≤ −2 was defined as the threshold for significant inhibition.

The integrative analysis of the three types of biological data (RNA-Seq, cell phenotypes, serum analytes) was performed using MOFA+ ^34^, a sparse Factor Analysis method. It provides latent variables which are linear combination of the most influential factors for explaining inter-patient variability across the three biological measurement modalities. The first component is presented and called integrative score here. The analyses of factors associated with CD177 protein concentration were performed using non parametric Wilcoxon test or Spearman correlation coefficient when appropriate. To look at the independent association of CD177 with ICU, a logistic regression for the prediction of hospitalization in ICU adjusted for age, sex, chronic cardiac disease, chronic pulmonary disease, diabetes was fitted. The analysis of repeated measurements of CD177 over time was done by using a linear mixed effect model adjusted for time from hospitalization and an interaction with survival outcome (death or recovery). The model included a random intercept and a random slope with an unstructured matrix for variance parameters. Predictions of marginal trajectories were performed. All analyses, if not stated otherwise, were performed using R software version 3.6.3 (R Core Team (2020)). R: A language and environment for statistical computing. R Foundation for Statistical Computing, Vienna, Austria. URL: https://www.R-project.org/

### Data availability

RNA sequencing data that support the findings of this study will be deposited in the Gene Expression Omnibus (GEO) repository. Other data will be provided as Source data files.

## Acknowledgments

We thank the patients who donated their blood. We thank F. Mentre, S. Tubiana, the French COVID cohort, and REACTing (REsearch & ACtion emergING infectious diseases) for cohort management. We thank the scientific advisory board of the French COVID-19 cohort composed of Dominique Costagliola, Astrid Vabret, Hervé Raoul, and Laurence Weiss. We thank Romain Levy for fruitful discussions.

## Author Contributions

YL and AW conceived and designed the study. MC, JFT, YY, LB, DB, GP, and MP participated in sample and clinical data collection. EF, MDe, MS, CLe, and PT performed the experiments. MD, MG, BH, and CLe analyzed the data. YL, RT, AW, HH, MS, BJH, and CL analyzed and interpreted the data. YL, RT, AW, and HH drafted the first version and wrote the final version of the manuscript. All authors approved the final version.

