## Supplementary Information for "CD177, a specific marker of neutrophil activation, is a hallmark of COVID-19 severity and death"

**Lévy *et al.***

**French Cohort Study Group**

Laurent ABEL (Inserm UMR 1163, Paris, France), Claire ANDREJAK (CHU Amiens, France), François ANGOULVANT (Hôpital Necker, Paris, France), Delphine BACHELET, Krishna BHAVSAR, Lila BOUADMA, Anissa CHAIR, Camille COUFFIGNAL, Charlene DA SILVEIRA, Marie-Pierre DEBRAY, Diane DESCAMPS, Xavier DUVAL, Philippine ELOY, Marina ESPOSITO-FARESE, Nadia ETTALHAOUI, Nathalie GAULT, Jade GHOSN, Isabelle GORENNE, Isabelle HOFFMANN, Ouifiya KAFIF, Sabrina KALI, Antoine KHALIL, Cédric LAOUÉNAN, Samira LARIBI, Minh LE, Quentin LE HINGRAT, François-Xavier LESCURE, Jean Christophe LUCET, France MENTRÉ, Jimmy Mullaert, Nathan PEIFFER-SMADJA, Gilles PEYTAVIN, Carine ROY, Marion SCHNEIDER, Nassima SI MOHAMMED, Lysa TAGHERSET, Coralie TARDIVON, Marie-Capucine TELLIER, Jean-François TIMSIT, Théo TRIOUX, Sarah TUBIANA, Benoit VISSEAUX, Yazdan YAZDANPANAH (Hôpital Bichat, Paris, France), Romain BASMACI, Olivier PICONE (Hôpital Louis Mourier, Colombes, France), Sylvie BEHILILL, Sylvie VAN DER WERF, Vincent ENOUF, Hugo MOUQUET (Pasteur Institute, Paris, France), Marine BELUZE (F-CRIN Partners Platform, Paris, France), Dehbia BENKERROU, Céline DORIVAL, François TÉOULÉ, Amina MEZIANE (Inserm UMR 1136, Paris, France), François BOMPART (Drugs for Neglected Diseases initiative, Geneva, Switzerland), Maude BOUSCAMBERT (Inserm UMR 1111, Lyon, France), Minerva CERVANTES-GONZALEZ, Eric d'ORTENZIO, Oriane PUÉCHAL, Caroline SEMAILLE (REACTing, Paris, France), Catherine CHIROUZE (CHRU Jean Minjoz, Besançon, France), Alexandra COELHO (Inserm UMR 1018, Paris, France), Sandrine COUFFIN-CADIERGUES, Hélène ESPEROU, Ikram HOUAS, Salma JAAFOURA, Aurélie PAPADOPOULOS (Inserm sponsor, Paris, France), Dominique DEPLANQUE (Hôpital Calmette, Lille, France), Mathilde DESVALLÉE, Coralie KHAN (Inserm UMR 1219, Bordeaux, France), Alpha DIALLO, Marie BARTOLI, Soizic LE MESTRE, Noémie MERCIER, Christelle PAUL, Ventzislava PETROV-SANCHEZ (ANRS, Paris, France), Alphonsine DIOUF, Alexandre HOCTIN, Marina MAMBERT (Inserm UMR 1018, Paris, France), François DUBOS (CHU Lille, France), Manuel ETIENNE (CHU Rouen, France), Alexandre GAYMARD (Inserm UMR 1111, Lyon, France), Tristan GIGANTE, Morgane GILG, Bénédicte ROSSIGNOL (F-CRIN INI-CRCT, Nancy, France), Jérémie GUEDJ, Hervé LE NAGARD, Guillaume LINGAS, Nadège NEANT (Inserm UMR 1137, Paris, France), Jean-Sébastien HULOT (Hôpital Européen Georges Pompidou, Paris, France), Florentia KAGUELIDOU, Justine PAGES (Hôpital Robert Debré, Paris, France), Yves LEVY, Aurélie WIEDEMANN (Vaccine Research Insitute (VRI), Inserm UMR 955, Créteil, France), Claire LEVY-MARCHAL (F-CRIN INI-CRCT, Paris, France), Bruno LINA, Manuel ROSA-CALATRAVA, Olivier TERRIER (Inserm UMR 1111, Lyon, France), Denis MALVY (CHU Bordeaux, France), Marion NORET (RENARCI, Annecy, France), Patrick ROSSIGNOL (CHU Nancy, France), Christelle TUAL, Aurélie VEISLINGER (Inserm CIC-1414, Rennes, France), Noémie VANEL (Hôpital la Timone, Marseille, France)

**Supplementary Table 1. Characteristics of COVID-19 patients described in the transcriptomic analysis**

| | No. | Group 1 (N = 10) | Group 2 (N = 16) | Group 3 (N = 18) |
|---|---|---|---|---|
| **Demographic characteristics** | | | | |
| Age - Median (IQR) - years | 44 | 57 (48 - 74) | 61 (51 - 66) | 61 (53 - 68) |
| Male sex - No./total No. (%) | 44 | 7/10 (70) | 14/16 (88) | 16/18 (89) |
| ICU or transfer to ICU or death - No /total No.(%) | 44 | 6/10 (60) | 13/16 (81) | 17/18 (94) |
| Outcome - No./total No. (%) | 44 | | | |
| Death | | 4/10 (40) | 4/16 (25) | 5/18 (28) |
| Discharge alive | | 6/10 (60) | 12/16 (75) | 13/18 (72) |
| **Median interval from first symptoms on admission (IQR)** | 44 | 7 (6 - 11) | 11 (10 - 14) | 13 (9 - 14) |
| **Comorbidities - No./total No. (%)** | | | | |
| Any | 44 | 2/10 (10) | 7/16 (44) | 4/18 (22) |
| Chronic cardiac disease | 44 | 4/10 (10) | 1/16 (6) | 2/18 (11) |
| Hypertension | 44 | 4/10 (40) | 2/16 (12) | 7/18 (39) |
| Chronic pulmonary disease | 44 | 0/10 (0) | 1/16 (6) | 2/18 (11) |
| Asthma | 44 | 1/10 (10) | 1/16 (6) | 0/18 (0) |
| Chronic kidney disease | 44 | 0/10 (0) | 0/16 (0) | 3/18 (17) |
| Chronic neurological disorder | 44 | 0/10 (0) | 1/16 (6) | 0/18 (0) |
| Obesity | 44 | 3/10 (30) | 5/16 (31) | 5/18 (28) |
| Diabetes | 44 | 1/10 (10) | 4/16 (25) | 6/18 (33) |
| **Smoking History - No./total No. (%)** | | | | |
| Smoking | 44 | 0/10 (0) | 3/16 (19) | 1/18 (6) |
| **Laboratory findings on admission - Median (IQR)** | | | | |
| Hemoglobin - g/dL | 44 | 13 (12 - 15) | 13 (12 - 14) | 12 (11 - 14) |
| WBC count - x10^9/L | 44 | 5 (5 - 6) | 6 (5 - 9) | 9 (6 - 13) |
| Platelet count - x10^9/L | 44 | 179 (156 - 198) | 188 (165 - 288) | 204 (115 - 359) |
| C-reactive protein (CRP) - mg/L | 44 | 61 (20 - 82) | 121 (60 - 196) | 152 (108 - 244) |
| Blood Urea Nitrogen (urea) - mmol/L | 44 | 6 (5 - 7) | 5 (3 - 10) | 8 (6 - 12) |
| **Symptoms on admission - No./total No. (%)** | | | | |
| Fever | 44 | 8/10 (80) | 14/16 (88) | 18/18 (100) |
| Cough | 42 | 5/8 (63) | 12/16 (75) | 13/18 (72) |
| Sore throat | 41 | 1/7 (14) | 1/16 (6) | 1/18 (6) |
| Wheezing | 39 | 1/8 (13) | 2/15 (13) | 0/16 (0) |
| Myalgia | 41 | 2/8 (25) | 8/16 (50) | 5/17 (29) |
| Arthralgia | 40 | 1/8 (13) | 2/15 (13) | 4/17 (24) |
| Fatigue | 42 | 2/8 (25) | 8/16 (50) | 8/18 (44) |
| Dyspnea | 42 | 6/8 (75) | 14/16 (88) | 15/18 (83) |
| Headache | 42 | 0/8 (0) | 5/16 (31) | 5/18 (28) |

| | | | | |
|---|---|---|---|---|
| Altered consciousness | 41 | 0/8 (0) | 0/15 (0) | 2/18 (11) |
| Abdominal pain | 38 | 0/8 (0) | 1/14 (7) | 3/16 (19) |
| Vomiting / nausea | 41 | 1/8 (13) | 2/15 (13) | 5/18 (28) |
| Diarrhea | 41 | 1/8 (13) | 4/15 (27) | 5/18 (28) |
| **Clinical characteristics on admission - Median (IQR)** | | | | |
| SOFA score | 23 | 3 (1 - 4) | 6 (4 - 7) | 6 (3 - 7) |
| SAPS2 | 24 | 37 (22 - 52) | 29 (28 - 38) | 46 (31 - 61) |
| Heart rate - beats per minute | 44 | 85 (80 - 92) | 87 (72 - 105) | 90 (75 - 103) |
| Respiratory rate - breaths per minute | 40 | 19 (18 - 20) | 27 (24 - 33) | 22 (19 - 33) |
| Systolic blood pressure - mmHg | 43 | 118 (108 -145) | 139 (114 -145) | 135 (121 -158) |
| Diastolic blood pressure - mmHg | 43 | 74 (65 - 80) | 78 (70 - 89) | 74 (71 - 82) |
| Oxygen saturation - percent | 44 | 95 (93 - 98) | 96 (91 - 97) | 93 (87 - 97) |
| Oxygen saturation on – No./total No. (%) | 44 | | | |
| Room air | | 4/10 (40) | 3/16 (19) | 7/18 (39) |
| Oxygen therapy | | 6/10 (60) | 13/16 (81) | 11/18 (61) |
| **Treatments - No./total No. (%)** | | | | |
| Antiviral | 43 | 5/10 (50) | 10/16 (62) | 14/17 (82) |
| Antibiotic | 43 | 4/10 (40) | 13/16 (81) | 12/17 (71) |
| Corticosteroids | 43 | 1/10 (10) | 8/16 (50) | 10/17 (59) |
| Antifungal | 43 | 0/10 (0) | 2/16 (12) | 3/17 (18) |
| Hydroxychloroquine | 43 | 0/10 (0) | 3/16 (19) | 3/17 (18) |

### Supplementary Table 2. All BCR related genes

| Genes | FDR | Log2 FC | Genes | FDR | Log2 FC | Genes | FDR | Log2 FC |
|---|---|---|---|---|---|---|---|---|
| IGHV1-24 | 0.030581621 | 5.407268 | IGHV3-73 | 0.0220482 | 2.725353 | IGHV1-17 | 0.01725323 | 2.231395 |
| IGLV3-10 | 0.026473188 | 4.282412 | IGLV3-9 | 0.0220482 | 2.712686 | IGLV2-28 | 0.04153633 | 2.201201 |
| IGLV3-25 | 0.012346539 | 3.90591 | IGLC1 | 0.01725323 | 2.701069 | IGKV6D-21 | 0.01234654 | 2.184745 |
| IGLV3-27 | 0.022048198 | 3.885845 | IGHJ4 | 0.01234654 | 2.669988 | IGLJ1 | 0.04473672 | 2.173331 |
| IGLV1-36 | 0.017253229 | 3.863016 | IGHJ1 | 0.01725323 | 2.6633 | IGLV8-61 | 0.01725323 | 2.173024 |
| IGLV3-1 | 0.00397524 | 3.837419 | IGHV4-61 | 0.0220482 | 2.656812 | IGKV1-6 | 0.0220482 | 2.168246 |
| IGLV4-69 | 0.017253229 | 3.739889 | IGKV3-15 | 0.0009382 | 2.651675 | IGKV2D-40 | 0.03438457 | 2.163532 |
| IGLV3-19 | 0.01061534 | 3.735987 | IGKV4-1 | 0.00028687 | 2.644773 | IGHV1OR15-4 | 0.03058162 | 2.151865 |
| IGHV1-18 | 0.017253229 | 3.667556 | IGHA1 | 0.00397524 | 2.634013 | IGHV3-71 | 0.0220482 | 2.089232 |
| IGHV3-21 | 0.017253229 | 3.587602 | IGKV1-27 | 0.04153633 | 2.63176 | IGKV1D-37 | 0.00746691 | 2.06909 |
| IGLV6-57 | 0.017253229 | 3.512432 | IGHV4-28 | 0.01725323 | 2.614677 | IGHV3-72 | 0.03058162 | 2.067323 |
| IGHV4-34 | 0.017253229 | 3.463972 | IGLV2-11 | 0.01234654 | 2.609628 | IGHV3-41 | 0.0220482 | 2.055012 |
| IGHV4-39 | 0.017253229 | 3.444361 | IGLV5-37 | 0.04153633 | 2.605725 | IGHG4 | 0.02647319 | 2.053045 |
| IGHV3-33 | 0.000286867 | 3.435046 | IGKV3-20 | 0.00028687 | 2.554925 | IGHV3-63 | 0.01725323 | 2.051185 |
| IGHV3-11 | 0.030581621 | 3.421957 | IGHV1-58 | 0.02647319 | 2.543716 | IGKV2D-28 | 0.01725323 | 2.018281 |
| IGHG3 | 0.000938197 | 3.407473 | IGKV1-5 | 0.00746691 | 2.511011 | IGKV3-7 | 0.01725323 | 1.963918 |
| IGHV1-46 | 0.022048198 | 3.39981 | IGKV2-29 | 0.01725323 | 2.488949 | IGHGP | 0.04786254 | 1.915987 |
| IGHV3-15 | 0.012346539 | 3.396577 | IGKV3-11 | 0.0220482 | 2.489234 | IGHV3-60 | 0.04153633 | 1.874073 |
| IGHV3-20 | 0.022048198 | 3.383072 | IGKV1D-33 | 0.04153633 | 2.460444 | IGKV2D-24 | 0.04786254 | 1.87279 |
| IGHV3-43 | 0.012346539 | 3.310178 | IGLJ3 | 0.0220482 | 2.421166 | IGHV3OR16-9 | 0.01725323 | 1.844052 |
| IGHV3-23 | 0.000286867 | 3.299521 | IGLV3-16 | 0.01725323 | 2.411455 | IGHG2 | 0.04473672 | 1.749984 |
| IGHV2-26 | 0.012346539 | 3.284427 | IGHV3-64 | 0.03058162 | 2.405988 | IGLV7-43 | 0.03810136 | 1.716134 |
| IGLV1-51 | 0.012346539 | 3.25168 | IGKV6-21 | 0.00053237 | 2.400168 | IGHV3-69-1 | 0.03810136 | 1.711671 |
| IGLC3 | 0.022048198 | 3.193759 | IGKV1-17 | 0.0220482 | 2.398496 | IGKV1OR2-108 | 0.04786254 | 1.670756 |
| IGLV1-44 | 0.017253229 | 3.081442 | IGHV3-74 | 0.00162353 | 2.393907 | IGKV1D-8 | 0.01725323 | 1.649229 |
| IGLJ2 | 0.012346539 | 3.054996 | IGLL1 | 0.70867098 | 0.2706642 | IGHJ3P | 0.00746691 | 1.609411 |
| IGHV3-13 | 0.026473188 | 3.03438 | IGHV3-19 | 0.01725323 | 2.376945 | IGKJ5 | 0.0220482 | 1.479375 |
| IGLC2 | 0.030581621 | 3.02678 | IGKV3D-15 | 0.01725323 | 2.367281 | IGHEP2 | 0.00162353 | 1.42042 |
| IGKV1-33 | 0.01061534 | 2.986319 | IGKV1D-39 | 0.01725323 | 2.354587 | IGHVIII-2-1 | 0.04473672 | -0.664831 |
| IGKV2D-30 | 0.04473672 | 2.971709 | IGHV4-55 | 0.01234654 | 2.353481 | | | |
| IGKV1D-16 | 0.01061534 | 2.964449 | IGKV1-9 | 0.02647319 | 2.31918 | | | |
| IGHV4-59 | 0.004544213 | 2.959443 | IGHJ2 | 0.04473672 | 2.307147 | | | |
| IGLV1-40 | 0.007466914 | 2.951791 | IGLV2-18 | 0.02647319 | 2.304288 | | | |
| IGHJ3 | 0.012346539 | 2.94855 | IGLV1-50 | 0.04153633 | 2.300749 | | | |
| IGKV3OR2-268 | 0.017253229 | 2.925686 | IGKC | 0.01725323 | 2.298358 | | | |
| IGHV5-51 | 0.007466914 | 2.914603 | IGHV1OR15-9 | 0.0220482 | 2.291757 | | | |
| IGLV10-54 | 0.022048198 | 2.911357 | IGLV7-46 | 0.02647319 | 2.273516 | | | |
| IGLV1-47 | 0.007466914 | 2.907312 | IGKV2-24 | 0.04786254 | 2.270762 | | | |
| IGHV3-49 | 0.012346539 | 2.905982 | IGLC6 | 0.04473672 | 2.264953 | | | |
| IGKV2-28 | 0.017253229 | 2.882124 | IGLV2-5 | 0.01061534 | 2.263166 | | | |
| IGLVI-70 | 0.034384566 | 2.859006 | IGKV3D-20 | 0.01234654 | 2.262339 | | | |
| IGHJ5 | 0.017253229 | 2.846426 | IGLV4-3 | 0.0220482 | 2.259118 | | | |
| IGLV2-23 | 0.012346539 | 2.835207 | IGHM | 0.02647319 | 2.257772 | | | |
| IGLV2-8 | 0.047862536 | 2.81604 | IGKV1-16 | 0.02647319 | 2.258257 | | | |
| IGLV2-14 | 0.000286867 | 2.802055 | IGHV1OR15-2 | 0.03058162 | 2.257497 | | | |
| IGLV9-49 | 0.012346539 | 2.800455 | IGKV5-2 | 0.0153988 | 2.242023 | | | |
| IGLV3-21 | 0.034384566 | 2.743388 | IGLV5-45 | 0.01725323 | 2.235904 | | | |

**Supplementary Table 3. Characteristics of healthy donors (HD) involved in the various assays**

| Assay | Immune cell phenotypes | Serum cytokine measurement | RNA-seq | Serum CD177 measurement |
|---|---|---|---|---|
| **n** | 18 | 5 | 10 | 16 |
| **Age – years - Median (IQR)** | 36 (30 - 45) | 30 (28 - 51) | 33 (29 - 38) | 26 (22 – 50) |
| **Male sex - nbr/total nbr (%)** | 16/18 (89) | 2/5 (40) | 10/10 (100) | 6/16 (38) |

**Supplementary Figure 1. Heatmap of standardized immune-cell subset phenotypic characterization in PBMCs.** The colors represent standardized expression values centered around 0, with variance equal to 1. HD: healthy donors (n = 18), COVID: COVID19 patients (n = 50). Each column represents a subject. Each line represents an immune-cell subset. The 18 healthy donors were combined to reduce the number of columns to 15, which corresponds to the number of healthy donors used for most of the immune-cell subsets. Only B cells were measured in 14 healthy donors, which explains the presence of missing data symbolized by black boxes.

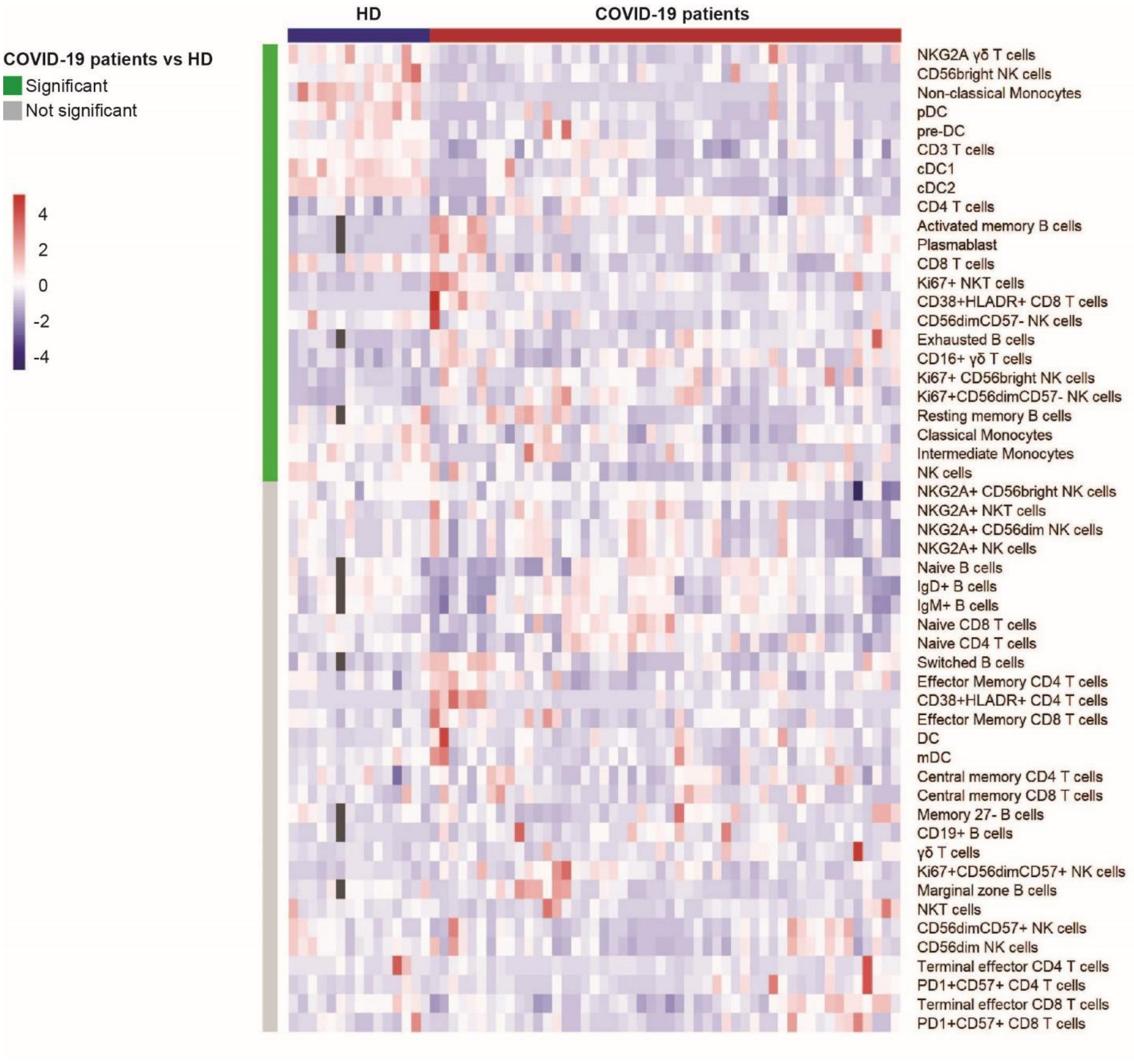

**Supplementary Figure 2. Quantification of soluble mediators differentially present in the serum of COVID19 patients.** Measurement of soluble serum mediators (pg/ml) from 5 HD and 33 COVID19 patients using the Bio-Plex 200 System$^{TM}$ (Bio-Rad). The differences between the two groups were evaluated using Wilcoxon rank sum statistical tests. The lower and upper boundaries of the box represent the 25% and 75% percentiles and the whiskers extend to the most extreme data point that is no more than 1.5 times the interquartile range away from the median. Median values (horizontal line in the boxplot) are shown.

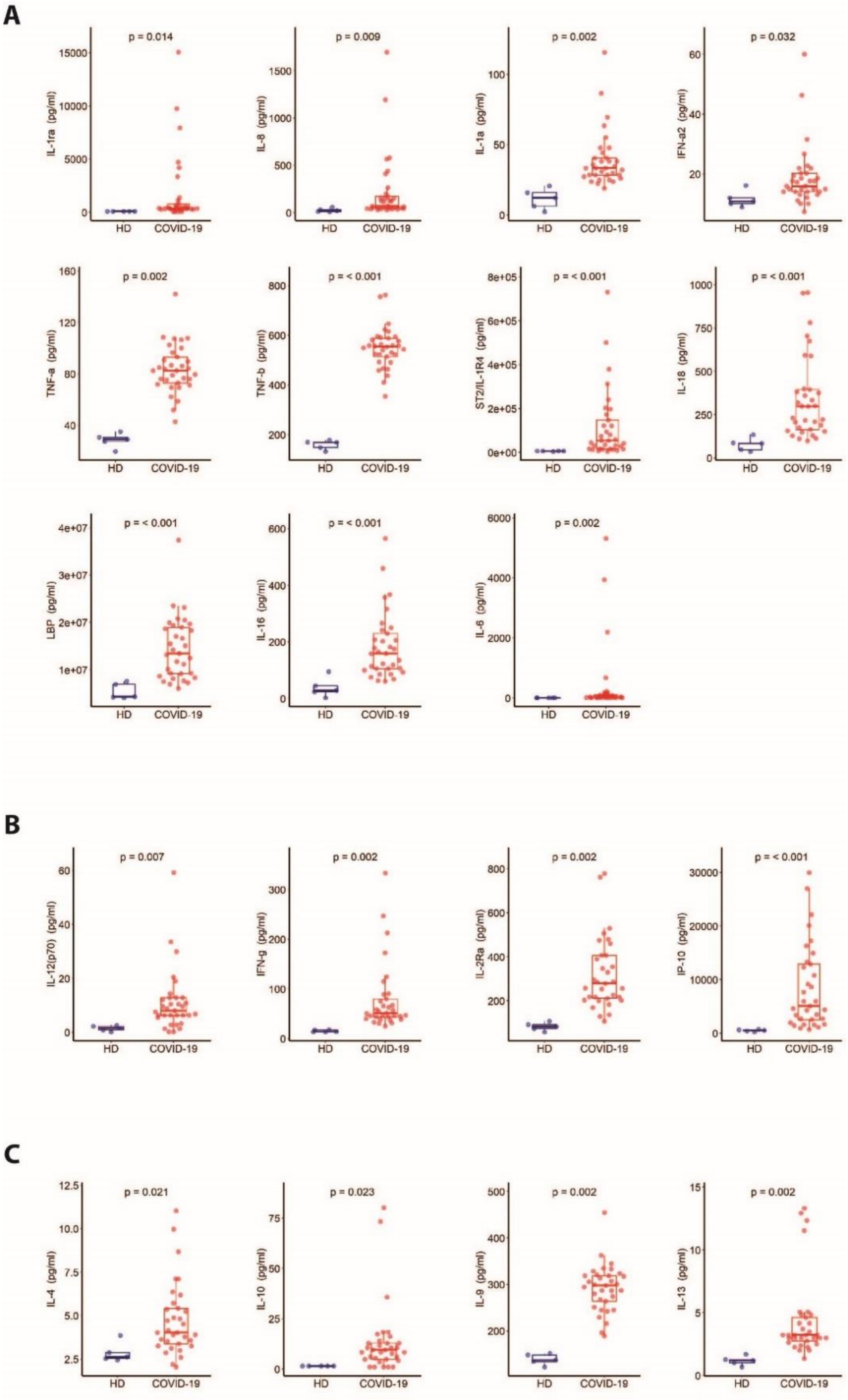

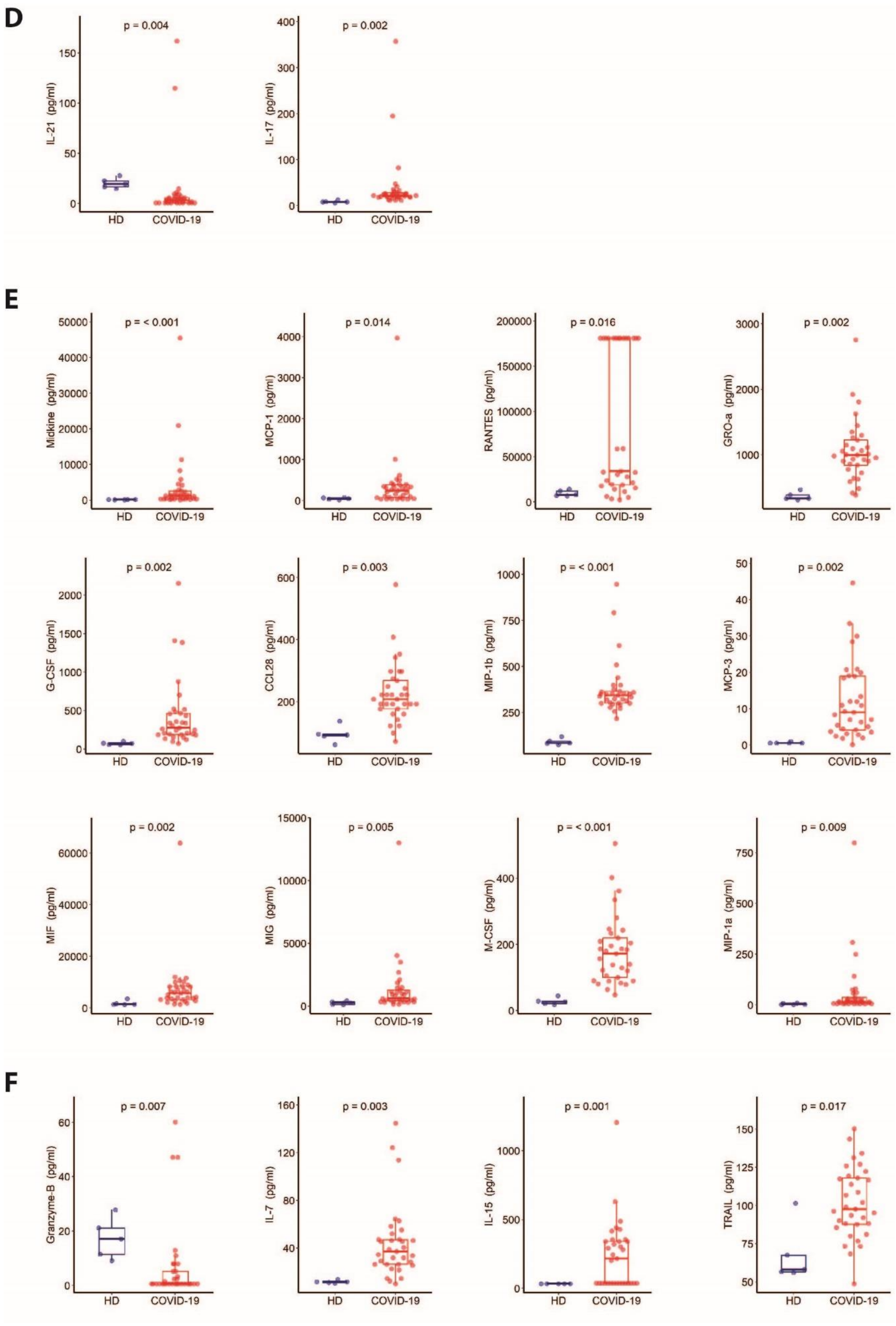
D
p = 0.004
IL-21 (pg/ml)
HD
COVID-19
p = 0.002
IL-17 (pg/ml)
HD
COVID-19
E
p = < 0.001
Midkine (pg/ml)
p = 0.014
MCP-1 (pg/ml)
p = 0.016
RANTES (pg/ml)
p = 0.002
GRO-a (pg/ml)
p = 0.002
G-CSF (pg/ml)
p = 0.003
CCL28 (pg/ml)
p = < 0.001
MIP-1b (pg/ml)
p = 0.002
MCP-3 (pg/ml)
p = 0.002
MIF (pg/ml)
p = 0.005
MIG (pg/ml)
p = < 0.001
M-CSF (pg/ml)
p = 0.009
MIP-1a (pg/ml)
F
p = 0.007
Granzyme-B (pg/ml)
p = 0.003
IL-7 (pg/ml)
p = 0.001
IL-15 (pg/ml)
p = 0.017
TRAIL (pg/ml)

**G**

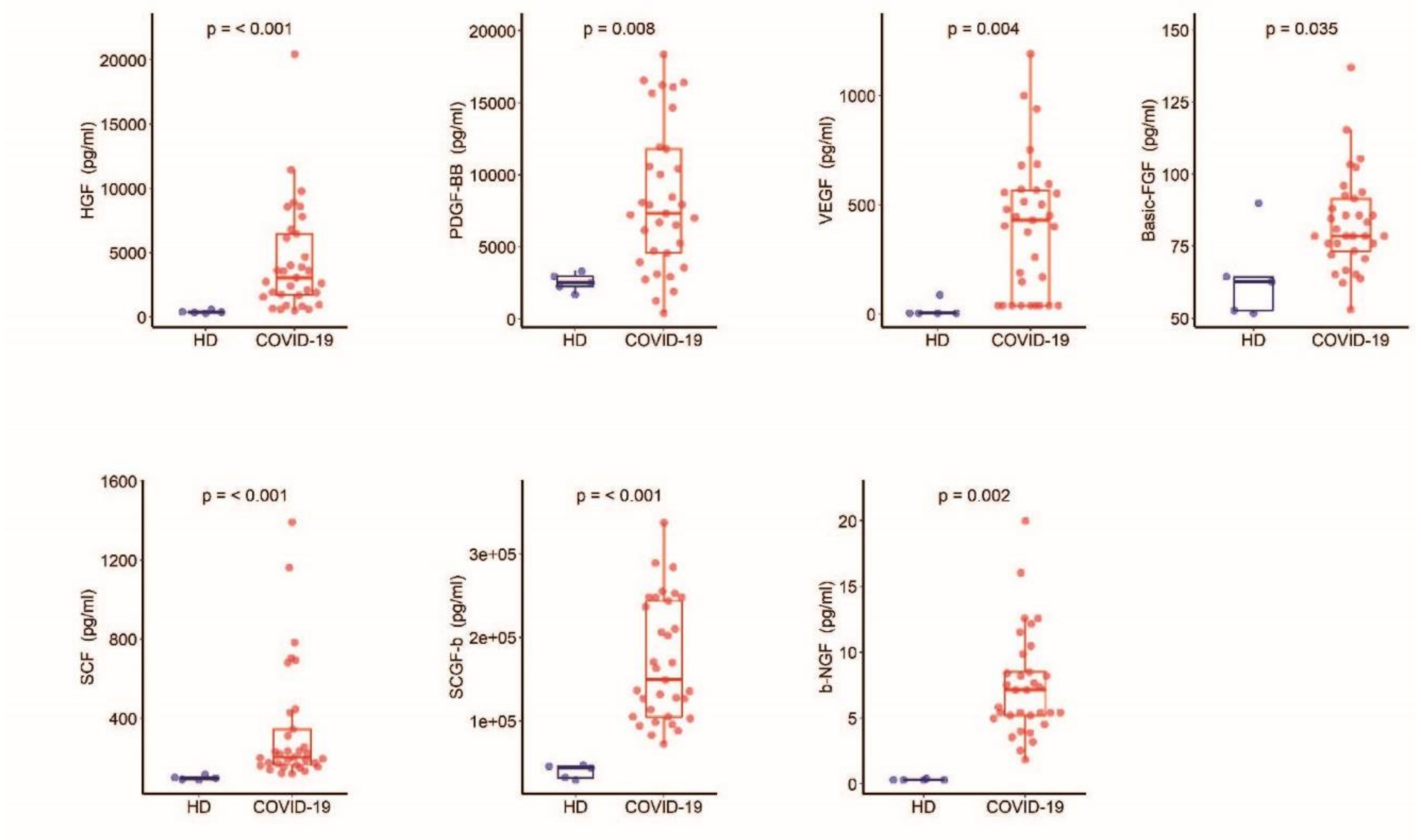

p = < 0.001
HGF (pg/ml)
20000
15000
10000
5000
0
HD
COVID-19
p = 0.008
PDGF-BB (pg/ml)
20000
15000
10000
5000
0
HD
COVID-19
p = 0.004
VEGF (pg/ml)
1000
500
0
HD
COVID-19
p = 0.035
Basic-FGF (pg/ml)
150
125
100
75
50
HD
COVID-19
p = < 0.001
SCF (pg/ml)
1600
1200
800
400
HD
COVID-19
p = < 0.001
SCGF-b (pg/ml)
3e+05
2e+05
1e+05
HD
COVID-19
p = 0.002
b-NGF (pg/ml)
20
15
10
5
0
HD
COVID-19

**Supplementary Figure 3. Top canonical signalling associated with DEG between COVID-19 and HD subjects.** Pathway analysis of DEG was processed using Ingenuity software and the main statistically significant FDRs are shown. **A** EIF2 signaling (FDR p- value = 1.34 $10^{-22}$). **B** mTOR signaling (FDR p-value = 3.84 $10^{-6}$). **C** ICOS-ICOSL signaling (FDR p-value = 3.2 $10^{-3}$).

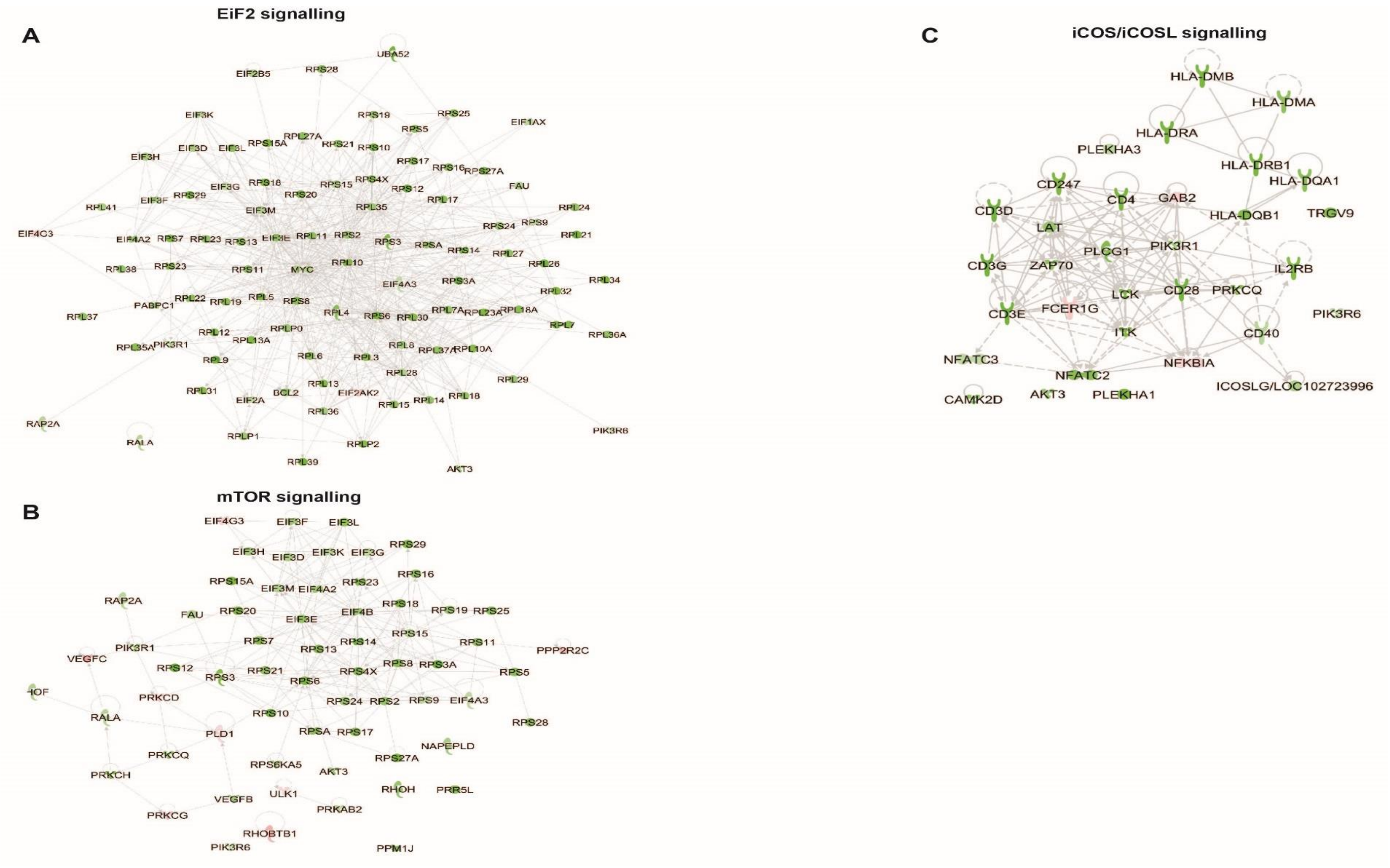

**Supplementary Figure 4. Integrative analysis from COVID-19 patients.** Integrative score from MOFA+ of the patient groups defined by the hierarchical clustering of the RNA-seq shown as in Figure 5A. The higher CD177 expression in log2 cpm normalized counts across groups is represented by the intensity of the color.

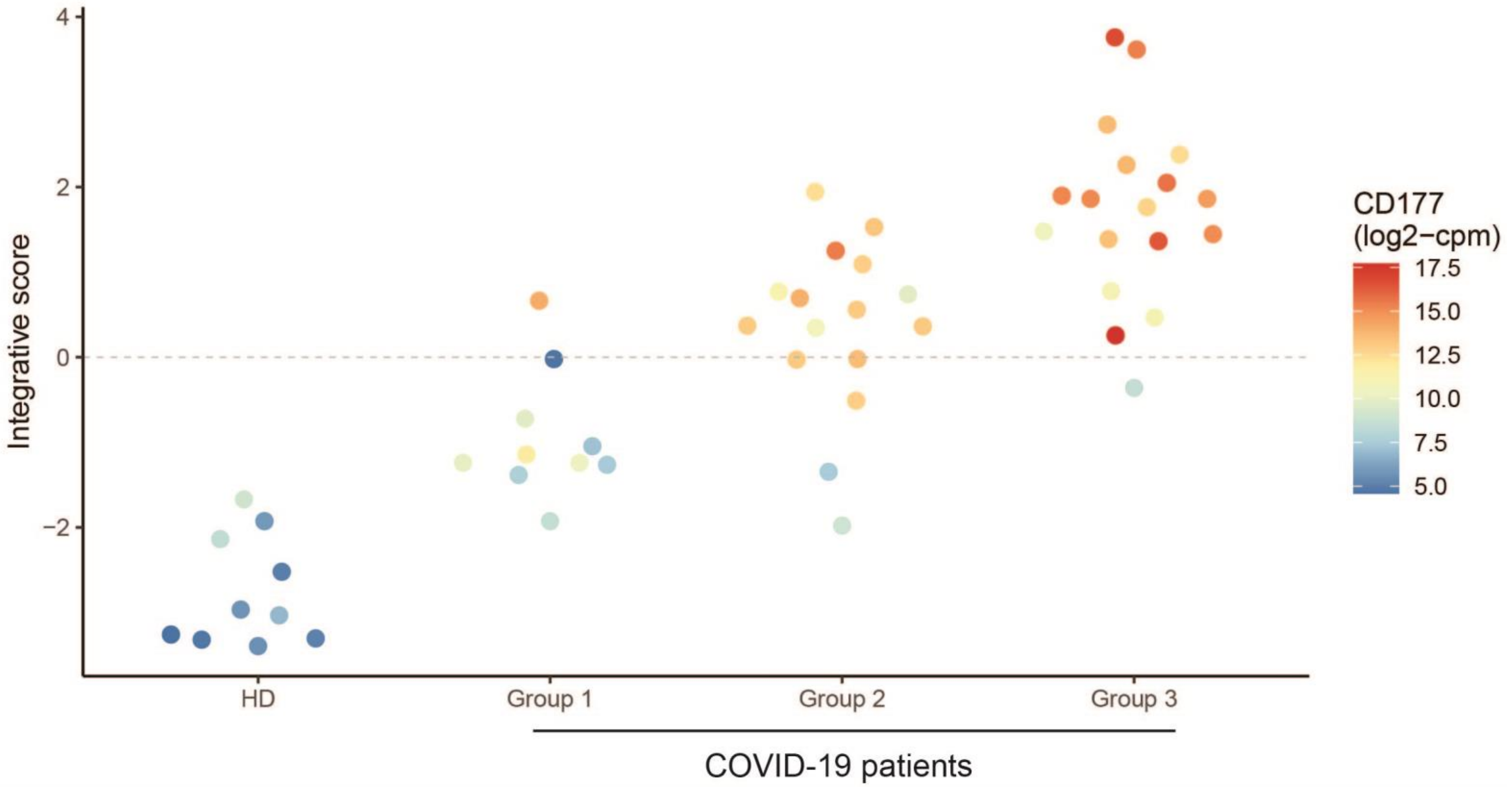

**Supplementary Figure 5.** Association between CD177 serum concentration and **A**. SOFA (n = 41) and **B**. SAPS2 (n=40) risk scores. The blue line represents the linear regression and the grey area is the 95% confidence interval.

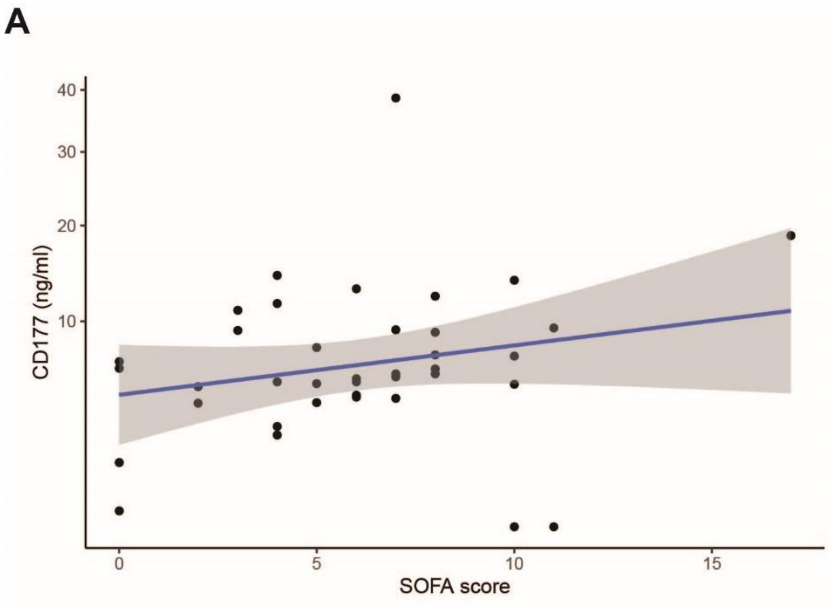


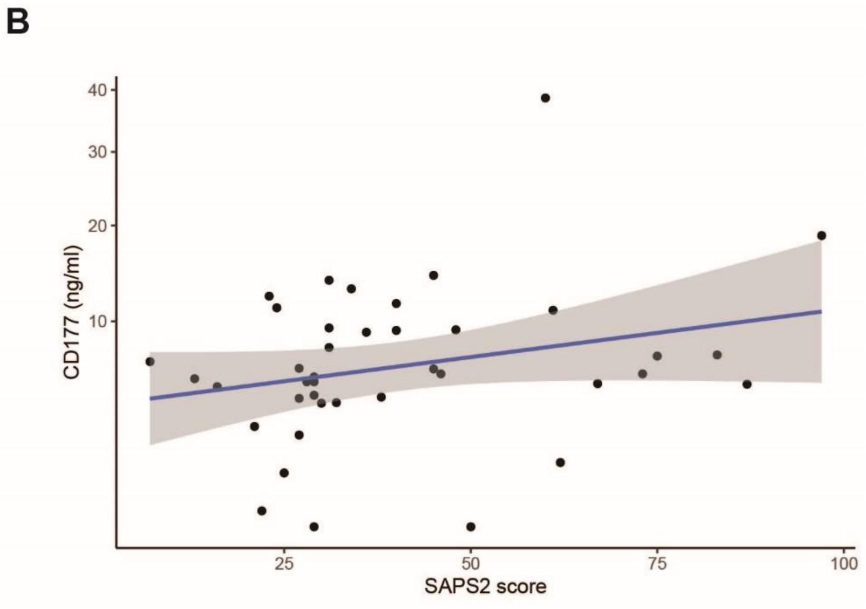
